# A single-molecule array urine test for tuberculosis: A case-control diagnostic accuracy study

**DOI:** 10.1101/2025.06.10.25329330

**Authors:** Tyler J. Dougan, Shira Roth, Liangxia Xie, Sydney D’Amaddio, David R. Walt

## Abstract

**Objectives:** Tuberculosis (TB) is one of the leading causes of death worldwide, even though it is curable using antibiotics. Most people who die of TB never begin treatment because diagnostics are insufficiently sensitive and accessible. We aimed to measure low-abundance biomarkers and diagnose TB in urine.

**Methods:** We developed and validated an ultrasensitive multiplex Single Molecule Array (Simoa) assay to detect TB in urine by measuring two TB biomarkers: lipoarabinomannan (LAM) and antigen 85B (Ag85B). Using antibodies that recognize different epitopes of LAM in a four-plex assay with three LAM and one Ag85B antibody pairs, we trained a model and demonstrated its performance in retrospective cohorts totaling 576 individuals from South Africa, Peru, Vietnam, and Cambodia, including a blinded test cohort (n=215).

**Results:** Our assay classifies samples with 98% specificity, 45% sensitivity overall, and 58% sensitivity among people living with the human immunodeficiency virus (HIV).

**Conclusions:** LAM concentrations measured in urine depend on the antibodies used to measure them. Our assay is more sensitive than the existing AlereLAM lateral flow test for TB in HIV-positive individuals, uses safe-to-use and accessible urine samples, and represents a first step towards an adjunctive diagnostic test to aid clinicians in starting treatment.

## BACKGROUND

Tuberculosis (TB) is the leading cause of death from a single infectious agent, killing over one million people each year [1]. It is caused by the bacillus *Mycobacterium tuberculosis* (*M. tb*) [1]. TB disease typically presents with a cough lasting more than three weeks, fever, weight loss, fatigue, and pain in the affected organ [2]. It usually affects the lungs (pulmonary TB, 85% of adult cases) but can also affect other sites in the body (extrapulmonary TB) [1]. TB disease is curable with six months to two years of antibiotics, but most people who die of TB never begin treatment [1]. In most countries’ TB programs, to begin treatment, individuals with TB must access diagnostic testing, be diagnosed, and be registered for treatment [1]. The proportion of people lost due to inaccessibility of testing, insensitive testing, and pre-treatment loss to follow-up varies between different health systems [1, 3, 4]. Many of the barriers that people face are structural, but addressing shortcomings in existing diagnostic tools could significantly reduce these barriers and potentially reduce TB morbidity [5].

About 60% of TB diagnoses are made with culture, smear microscopy, and nucleic acid amplification tests (NAATs) [1]. These tests provide microbiological confirmation of TB, which is essential because it is specific for the pathogen, authoritative to both patients and health systems, and necessary for drug resistance testing [1]. However, they rely on sputum samples, which have much higher loads of detectable bacilli than other specimen types. Sputum from people with pulmonary TB is highly infectious, and healthcare workers who collect or process sputum contract TB at much higher rates than the general population [6]. Some patients with TB symptoms (especially children, people living with HIV, and people with extrapulmonary TB) cannot produce enough sputum for testing [7]. The requirements for conducting these procedures safely are a major driver of the cost of TB diagnostic clinics and laboratories, limiting their expansion and accessibility [8]. Therefore, unfortunately, access to these TB testing is limited where it is most needed.

Some people reach testing centers but are not accurately diagnosed with TB because existing testing methods are insufficiently sensitive. These undiagnosed cases have been estimated to be 17% in India and 14% in South Africa in 2013, with considerable uncertainty [3, 4]. Culture is the most authoritative microbiologic test for diagnosing TB disease; its sensitivity is usually estimated at around 80%, but because it is used as a reference standard, other tests’ accuracies are usually reported relative to culture, which is then defined as 100% [9]. It takes an average of 13 days in liquid media or 26 days in solid media in a centralized laboratory with biosafety level 3 [10]. Sputum smear microscopy, used more widely for initial diagnosis, has a sensitivity of only 52% [11]. However, it takes only a few hours of processing time in a much simpler laboratory and can return results to patients in 1–3 days [12]. NAATs, including the Cepheid Xpert MTB/RIF (approved by the WHO as rapid diagnostic test) and Molbio Truenat MTB, use the polymerase chain reaction (PCR) to amplify genetic material [7]. Both tests take about two hours to complete, involve two simple hands-on steps, and can test for resistance to the first-line drug rifampicin [13]. Xpert is more sensitive and widely used, while Truenat is more portable, even able to run on battery power [14]. The second-generation Xpert Ultra test has a sensitivity of 91%, including 99% in smear-positive TB and 78% in smear-negative TB, with 96% specificity [15]. Truenat has been tested less than Xpert, but pooled estimates from two studies of its second generation, Truenat Plus, would assign it 98% and 53% sensitivity in smear-positive and smear-negative TB, respectively, and 96% specificity [14, 16]. These NAATs are less sensitive in children and people with extrapulmonary TB or comorbid HIV [7].

In summary, the existing diagnostic tools are not sufficient to stop TB. In 2023, 38% of diagnoses were made based on clinical signs, symptoms, and chest radiography, which do not provide microbiological confirmation [1]. Bacteriological culture cannot be a primary test because it takes weeks to return a result [10]. Smear microscopy fails to detect about half of TB cases [11]. Xpert and other molecular tests are faster and more sensitive, but still rely on sputum, which is highly infectious and not always available. The World Health Organization (WHO) has approved one non-sputum-based test: the Abbott (formerly Alere) Determine^TM^TB LAM Ag test (AlereLAM) [7]. AlereLAM is a lateral flow assay for the *M. tb* antigen lipoarabinomannan (LAM) in urine that gives results in 25 minutes [2]. However, it is only recommended for use in people with HIV, who comprise 6% of TB cases; even then, it is only 42% sensitive and 91% specific [2, 7].

In 2014, the WHO released target product profiles (TPPs) for four urgently needed TB diagnostics, including a rapid biomarker-based non-sputum-based test for detecting TB [17]. Ten years later, this unmet need was updated with a TPP on a rapid test for detecting *M. tuberculosis* at the peripheral level [18]. The TPP specifies an ideal diagnostic test that should involve three or fewer manual steps, deliver results in one hour, process at least eight samples per day, and have built-in calibration and controls [18]. The TPP’s guidelines on price per test and clinical sensitivity are stratified by setting: point-of-care (POC; instrument-free tests usable without training like AlereLAM, ≤$4 and ≥65% sensitive), near-POC (battery-operated lab-free tests like Truenat, ≤$6 and ≥75% sensitive), and low-complexity (clinical or microscopy laboratory device like Xpert, ≤$8 and ≥80% sensitive) [18]. The TPP also sets a minimum specificity of 98% to rule out other TB-like illnesses and latent or prior TB [18]. Urinary biomarkers would be ideal for this TPP, as urine is plentiful, safe to handle, and simple to collect from adults and children without generating hazardous bioaerosols [19]. The TB biomarker supported by the most evidence is lipoarabinomannan (LAM), the glycolipid detected by the AlereLAM assay [20]. LAM is released from metabolically active or degrading bacilli in the lungs or other sites of TB disease. Then, it is passed into the bloodstream, where it associates with lipoproteins. LAM is filtered in the glomeruli and enters the urine. It is detectable in sputum and urine but appears at much lower levels in serum or plasma unless aggressive extraction procedures are used [19]. LAM levels in urine vary widely, but they tend to be highest in people living with HIV and people with more advanced TB, disseminated TB, or renal TB [19]. LAM is non-covalently attached to the mycobacterial plasma membrane via a glyco-phospholipid anchor, one of its three structural domains. The phospholipid anchor is attached to a carbohydrate mannose core, conserved across all mycobacterial species. From that core, an arabinan carbohydrate domain branches with variable side chains and mannose capping [19]. The different forms of mannose capping of the arabinose side chains give rise to the diversity of LAM molecules [19].

LAM has an average molecular weight of 17 kDa, and epitopes are repeated multiple times [21]. LAM can be detected using different antibodies to different epitopes [22]. For example, the S4-20 (Otsuka Pharmaceutical) monoclonal antibody detects Man2 or Man3 caps further modified with a 5-methylthio-xylofuranose (MTX) cap [20]. FIND28 (Foundation for Innovative New Diagnostics (FIND)) detects arabinose6 (Ara6) cap with or without any Man cap [20]. A194-01 (Rutgers University) monoclonal antibody detects Ara4, Ara6, with or without Man1 cap [21]. G3 (Otsuka Pharmaceutical) monoclonal antibody detects Man2 and Man3 cap epitopes [20, 22]. Differences in the reactivity of antibodies to LAM from culture and urine have been observed, suggesting that urine LAM structure differs from native LAM released from bacteria [20]. While some studies have explored these antigenic differences, they remain poorly understood [23].

*M. tb* also has a number of promising protein biomarkers, including the diacylglycerol acyltransferase/mycolyltransferase antigen 85 complex of fibronectin-binding proteins (Ag85, *fbpB*) [24] Ag85B is one of the most antigenic proteins of *M. tb* [25].

There is a strong association between analytical sensitivity (limit of detection, LOD) and clinical sensitivity in LAM tests. For example, AlereLAM’s LOD is not reported, but it is likely around 1 ng/mL [19] and its clinical sensitivity is low (42% sensitivity [2]),. Paris et al. (2017) showed that LAM was detectable in urine from HIV-negative people with TB using assay with an LOD of 14 pg/mL [26]. Sigal et al. (2018) compared many antibody pairs in a sandwich immunoassay format (Meso Scale Discovery; this assay is sometimes referred to as MSD or EclLAM due to its electrochemiluminescent readout) [20]. They found that two pairs of monoclonal antibodies (S4-20/A194-01 and FIND28/A194-01) yielded assays with very low LODs (6 and 11 pg/mL, respectively) and high clinical sensitivity in smear-positive TB [20]. The first of these pairs was used to develop a lateral flow assay, Fujifilm SILVAMP TB LAM (FujiLAM), which stalled after studies reported lot-to-lot variability [27, 28]. As the reported concentrations of LAM are in the pg/mL order of magnitude and the current AlereLAM and EcLAM are not sensitive enough, we aimed to develop an ultrasensitive assay for LAM.

Many tools can test for proteins and similar macromolecules, but single-molecule arrays (Simoa) are ideal for detecting TB in diverse patients because they can quantify analytes present at very low concentrations [29]. Simoa is based on sandwich enzyme-linked immunosorbent assays (ELISAs), which use the selectivity of a pair of antibodies to quantify an analyte macromolecule [29]. In Simoa, these reactions take place on beads, so that the signal from a single binding event can be detected. Paramagnetic beads are coated with a capture antibody, and these beads are added to the solution to be tested. Because hundreds of thousands of beads are added to a relatively small volume, mass transport to the bead surface is rapid. For low-abundance measurements (fM–aM), there are many more beads than molecules, so that after binding most beads contain no molecules and some contain a single bound molecule. Beads are washed to remove interfering molecules, the detection antibody is added, and an enzyme is bound to the detection antibody. After washing, the only enzymes left are those attached to a bead with an analyte molecule bound. The beads are loaded into wells with the enzyme’s substrate, which the enzyme converts into a fluorescent product. A camera images and counts the number of fluorescent wells (meaning they have a protein molecule bound) and the total number of wells containing a bead. These values are used to calculate the average enzyme per bead (AEB), which is converted back to a concentration by generating a calibration curve with different dilutions of a purified form of the analyte. Whereas traditional ELISA uses a bulk measurement of total fluorescence, Simoa isolates beads into individual wells, and because there are many more beads than analyte molecules, the number of molecules can be calculated from the number of fluorescent beads using Poisson statistics.

A Simoa assay was developed to measure LAM in serum with high analytical sensitivity but low clinical sensitivity, likely due to shielding by lipoproteins as discussed above [30]. In another study, Simoa was used to measure four cytokines and immunoglobulins to one *M. tb* protein, Ag85B, in serum and plasma [31]. Yet, higher sensitivity and specificity are needed for a diagnostic test.

Here, we report developing and validating a new robust ultrasensitive Simoa assay to diagnose TB by measuring both LAM and Ag85B in urine. We demonstrate its sensitivity and specificity using cohorts totaling over 550 individuals. We exploit antibodies that recognize different epitopes on LAM antigen to develop an ultrasensitive four-plex classification assay with three LAM and one Ag85B antibody pairs. The assay identifies Ag85B and LAM antigens in urine and classifies the samples as TB positive or negative with 98% specificity, 45% sensitivity overall, and 58% sensitivity among people living with HIV. Our assay is more sensitive than AlereLAM, and although our assay is less sensitive than Xpert Ultra and Truenat Plus, it uses safe and accessible urine samples (non-sputum-based). Our assay is a first step towards a diagnostic test that may supplement existing diagnostics and aid clinicians in deciding whether to start TB treatment.

## METHODS

### 1. Antibodies and standards

The following reagents were obtained through BEI Resources, NIAID, NIH: *Mycobacterium tuberculosis*, Strain H37Rv, Purified Lipoarabinomannan (LAM), NR-14848 and Ag85B (Gene Rv1886c), Purified Native Protein from *Mycobacterium tuberculosis*, Strain H37Rv, NR-53526. FIND28 antibody was obtained from the Foundation for Innovative New Diagnostics (FIND; Geneva, Switzerland). G3 and S4-20 antibodies were kindly provided by Otsuka Pharmaceutical Co., (Tokyo, Japan). A194-01 IgM detection antibody for LAM was provided by Rutgers University (Newark, NJ). Ag85B capture (182λ) and detection (149κ) antibodies were obtained from AbCellera Biologics (Vancouver, Canada) under material transfer agreements. GenScript was contracted to produce large batches of antibodies using sequence information provided by AbCellera Biologics under non-disclosure agreements.

### 2. Study design

Assays were developed for 11 *M. tb* antigens in urine and tested in a discovery cohort; two (LAM and Ag85B) were selected for further optimization. After development and validation, this multiplexed ultrasensitive assay for LAM and Ag85B was evaluated in a retrospective case-control study to build and evaluate a diagnostic model for TB.

This study was exempted by Partners Human Research Committee (Protocol #2017P001447). All urine samples were provided by the FIND TB sample repository. FIND collected these samples under institutional review board (IRB)/independent ethics committee (IEC) approved studies in participating countries. Urine samples were collected from adult subjects with symptoms suggestive of pulmonary TB in South Africa, Peru, Vietnam, and Cambodia between June 2012 and February 2019, and all samples were collected prior to medical intervention. All clinical data were anonymized, and barcodes were used to access pertinent information. FIND uses standardized protocols for collecting and processing samples, which were reported previously in detail [20]. Obtained subject samples were categorized based on microbiological assessment. TB-positive individuals were patients with at least one positive culture result. Individuals categorized as non-TB were smear-negative and culture-negative from all sputum samples, negative for GeneXpert if tested, and their symptoms were improved or recovered in the absence of TB treatment at the follow-up symptom screening. The subjects were also classified as HIV-positive or HIV-negative on the basis of HIV rapid tests.

The study consisted of three cohorts: a model-building (training) cohort of 120 participants, a validation cohort of 251 participants, and blinded test cohort of 217 participants (Table 1). Samples were selected retrospectively from FIND’s specimen bank based on TB category (S+C+TB, S–C+TB, clinically diagnosed TB, non-TB/non-LTBI, non-TB/LTBI, and likely subclinical TB) and HIV status. Cohorts were procured sequentially: (1) a convenience cohort of 100 samples to establish performance; (2) a validation cohort of 258 procured based on a power calculation to demonstrate 90% sensitivity with 5.2% margin of error [32]; (3) an added 40 smear-negative samples distributed between the above two cohorts, which otherwise had none; and (4) a blinded test cohort of 244 samples, limited by the number of smear-negative and culture-negative samples available. (The totals in Table 1 differ because they exclude cases where multiple aliquots of the same sample were shipped by FIND in different cohorts. Results from these duplicate aliquots were combined and assigned to the earliest cohort in which the sample appeared, in order to preserve blinding.)

**Table 1.**
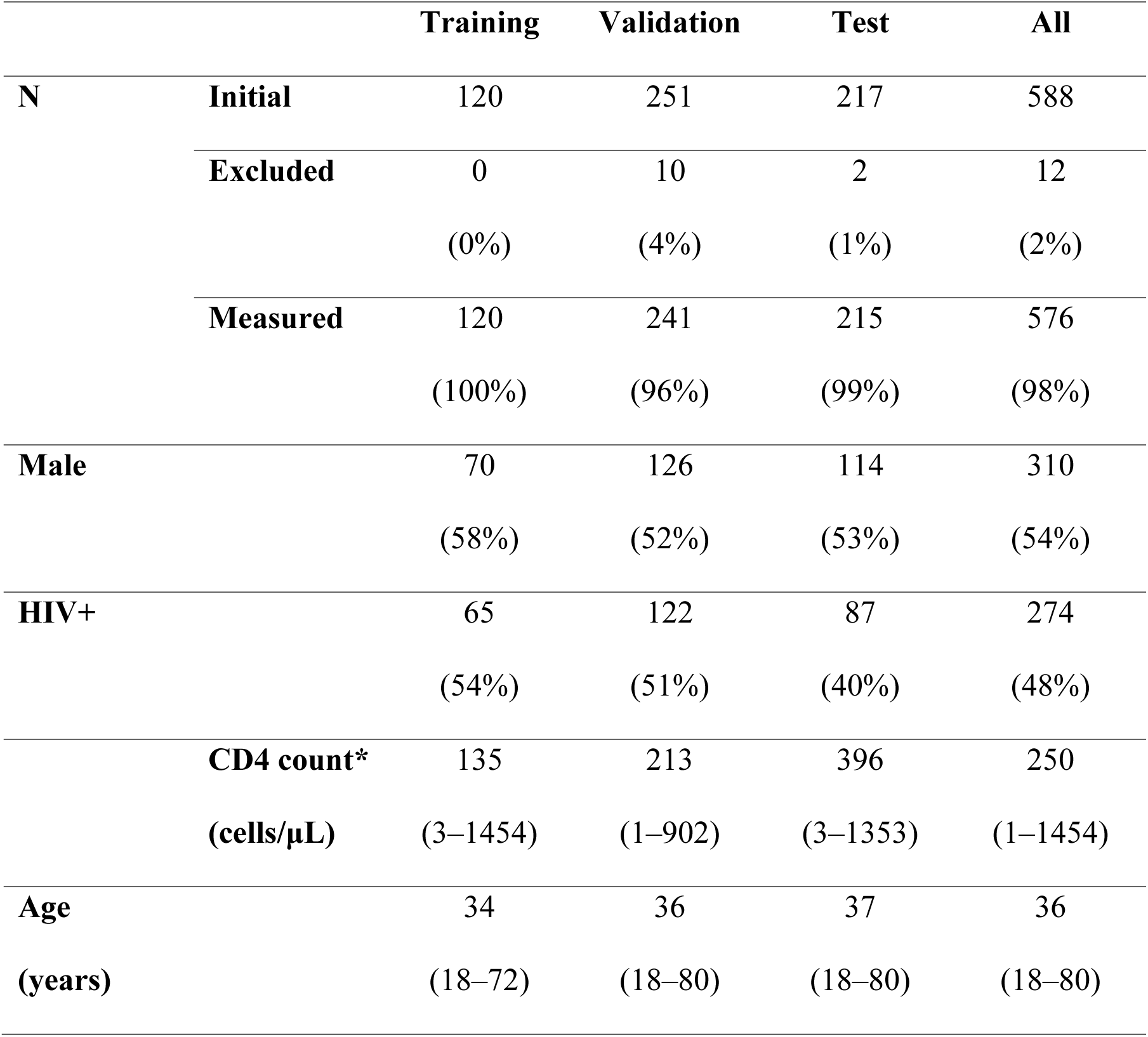
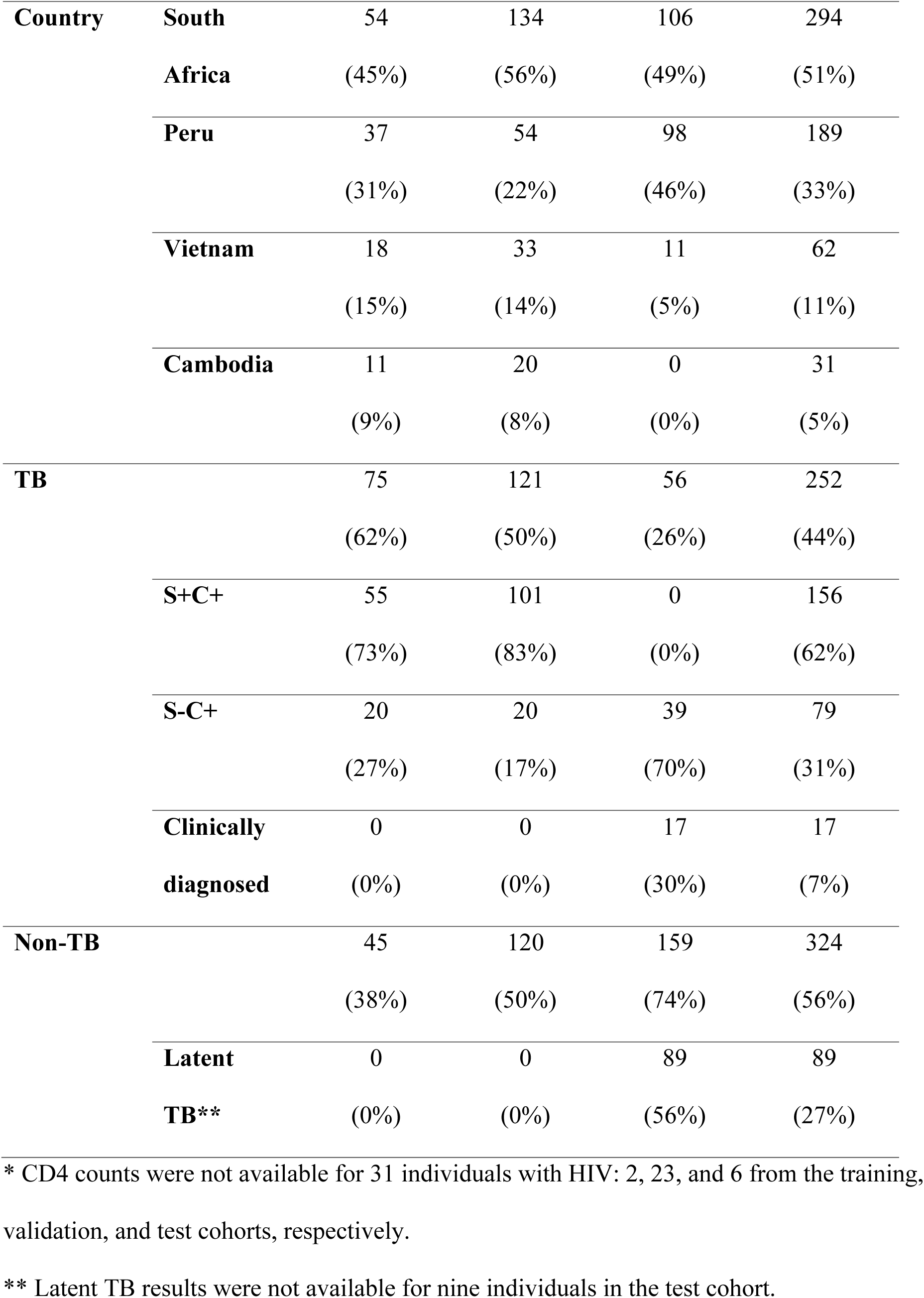
Clinical characteristics. Characteristics of the three cohorts. Values are given as *N* (%)

Samples were labeled only with barcodes that contained no clinical information. Investigators had access to the clinical data for the first three cohorts but avoided looking at it until the appropriate stage of data analysis after conducting Simoa assays. Investigators knew the numbers of samples in each TB and HIV category in the blinded test cohort, but nothing about the individual samples, and neither the barcodes nor the order nor any other features of the sample tubes suggested the eventual results. Simoa and AlereLAM assays were conducted independently on the blinded test cohort; a model trained on the prior cohorts was used to predict diagnoses in the test cohort using the Simoa results without seeing the AlereLAM results or any clinical information. Finally, the Simoa concentrations, model predictions, and AlereLAM results in the test cohort were submitted to FIND before FIND returned the clinical results and other information about this cohort.

Three replicates were measured from one urine sample for each individual. When Simoa image analysis returned an error (as in 4% of replicates) or the three replicates had coefficients of variation above 20%, additional replicates were run. The reported concentrations for each sample are the medians of all replicates measured. Twelve samples were excluded because they repeatedly failed Simoa runs; most of these had grossly visible anomalies. No other samples were excluded; no outliers were removed or adjusted.

### 3. Single Molecule Array assays

Simoa assays were performed using the Simoa HD-X Analyzer (Quanterix, Billerica, MA). Simoa is based on sandwich ELISAs, which use the selectivity of a pair of antibodies to quantify an analyte macromolecule. Bead conjugation, detector biotinylation, and Simoa assays were performed according to modified versions of protocols published previously [29].

*Bead conjugation*—Capture antibodies were reconstituted or buffer exchanged into 50 mM 2-(N-morpholino)ethanesulfonic acid (MES) buffer (Quanterix, Billerica, MA) via three washes through a 50 kDa Amicon Ultra-0.5 centrifugal filter according to the manufacturer’s instructions. Recovered antibodies were diluted to 0.2 mg/mL in 50 mM MES. A volume of 2.5 μm carboxylated paramagnetic beads (Quanterix, Billerica, MA) containing 1.4 × 109 beads per mL of diluted antibody was transferred to a new microcentrifuge tube. Beads were washed by placing the microcentrifuge tube on a magnetic separator, waiting for the beads to collect, aspirating the supernatant, removing the tube from the magnetic separator, adding fresh buffer, and vortexing to mix. Beads were washed three times in Bead Wash Buffer (Phosphate-buffered saline (PBS) with Tween 20, Quanterix, Billerica, MA) and three times in 50 mM MES (Quanterix, Billerica, MA). Carboxyl groups on the beads’ surfaces were then activated by incubating in 0.3 mg/mL 1-Ethyl-3-(3-dimethylaminopropyl)carbodiimide (EDC) (Thermo Fisher) in 50 mM MES for 30 minutes at 4 °C, with gentle shaking to keep them in suspension. Beads were washed once with 50 mM MES and then incubated with the 0.2 mg/mL capture antibody for 2 hours at 4 °C, with gentle shaking to keep them in suspension. Beads were washed twice in Bead Wash Buffer and blocked by incubating in Bead Blocking Buffer (PBS with Bovine Serum Albumin (BSA), Quanterix, Billerica, MA) for 45 min at room temperature or overnight at 4 °C. Finally, beads were washed once with Bead Wash Buffer, once with Bead Diluent (Tris-buffered saline (TBS) with Tween 20 and BSA, Quanterix, Billerica, MA), resuspended in Bead Diluent, and counted using a Coulter Counter. The typical batch volume was 300 μL (4.2 × 108 beads), but volumes were adjusted proportionally for larger and smaller batches.

*Detector biotinylation*—Detection antibodies were reconstituted or buffer exchanged into PBS (Quanterix, Billerica, MA) via three washes through a 50 kDa Amicon Ultra-0.5 centrifugal filter according to the manufacturer’s instructions. Recovered antibodies were diluted to 1 mg/mL in PBS. NHS-PEG4-biotin (N-Hydroxysuccinimide (NHS)-polyethylene glycol 4 (PEG4)-biotin,Thermo Fisher) was diluted to 8.9 mM in deionized water (2 mg NHS-PEG4-biotin / 383 μL water) and added to detection antibody at a concentration of 0.267 mM (0.157 mg/mL), corresponding to a 40-fold molar ratio of biotin to antibody monomer. In a typical batch, 3 μL of 8.9 mM biotin were added to 100 μL of antibody. After mixing, the solution was incubated for 30 minutes at room temperature. Excess biotin was removed in three washes of PBS through a 50 kDa Amicon Ultra-0.5 centrifugal filter according to the manufacturer’s instructions, and the final antibody concentration was determined with a NanoDrop spectrophotometer.

*Sample preparation*—Frozen urine samples were thawed in a room-temperature water bath, inverted to resuspend sediment, and pipette mixed with equal volumes of sample diluent in protein low-binding tubes (Eppendorf). The sample diluent consisted of 2X PBS (pH 7.4, 4 mM phosphate ion, Gibco) with 4% BSA (heat shock fraction, MilliporeSigma), 10 mM Ethylenediaminetetraacetic acid (EDTA) (Thermo Fisher), and 0.06% ProClin 300 (MilliporeSigma). Either 210 μL (for one replicate) or 390 μL (for two replicates) of diluted sample was transferred to a 96-well plate (Quanterix) and loaded into the HD-X. At least three replicates were measured for all samples. A calibration curve, consisting of serial dilutions of purified native LAM and Ag85B (BEI Resources) in calibrator diluent (1X PBS with 2% BSA, 5 mM EDTA, and 0.03% ProClin 300), was measured with each batch of samples.

*Simoa assay*—A Quanterix HD-X Analyzer automatically performed the 3-step Simoa assay at room temperature. First, 170 μL of each diluted sample or calibrator was combined with 25 μL of bead reagent in a reaction cuvette and incubated for 30 minutes with shaking to keep the beads in suspension. The bead reagent consisted of 5 × 106 beads/mL of each of the four plexes of beads (one for Ag85B and three for LAM, as shown in Figure 1) in a bead diluent of 50 mM TBS with 2% BSA, 0.1% Tergitol (NP-40, MilliporeSigma), and 0.03% ProClin 300. The beads were drawn to the side of the cuvette with a magnet and washed several times with System Wash Buffer 1 (5X PBS with Tween 20, Quanterix) to remove unbound sample. In the second step, beads were incubated for 5 minutes in 100 μL of detector reagent, consisting of 0.6 μg/mL of biotinylated Ag85B antibody 149 and 0.1 μg/mL of biotinylated LAM antibody A194-01 IgM in the detector diluent: PBS with 2% BSA, 5 mM EDTA, 0.1% Tergitol, and 0.03% ProClin 300. In the third step, after several washes with System Wash Buffer 1 to remove unbound detection antibodies, the beads were incubated for 5 minutes in 100 μL of streptavidin-β-galactosidase (SBG) reagent, consisting of 100 pM SBG in SBG Diluent (PBS with BSA, Tween 20, ProClin 300, and an interference blocker, Quanterix). Finally, excess SBG was washed away using Wash Buffer 1 followed by PBS (Quanterix) to remove Tween 20, and the beads were resuspended in 25 μL of RGP and loaded into an array of femtoliter-sized wells. The wells were sealed with oil and imaged using a fluorescence microscope. In wells containing beads with SBG bound (“on” beads), the SBG converted RGP to a fluorescent product, resorufin, that was visible in the 574/615 nm image.

**Figure 1.**
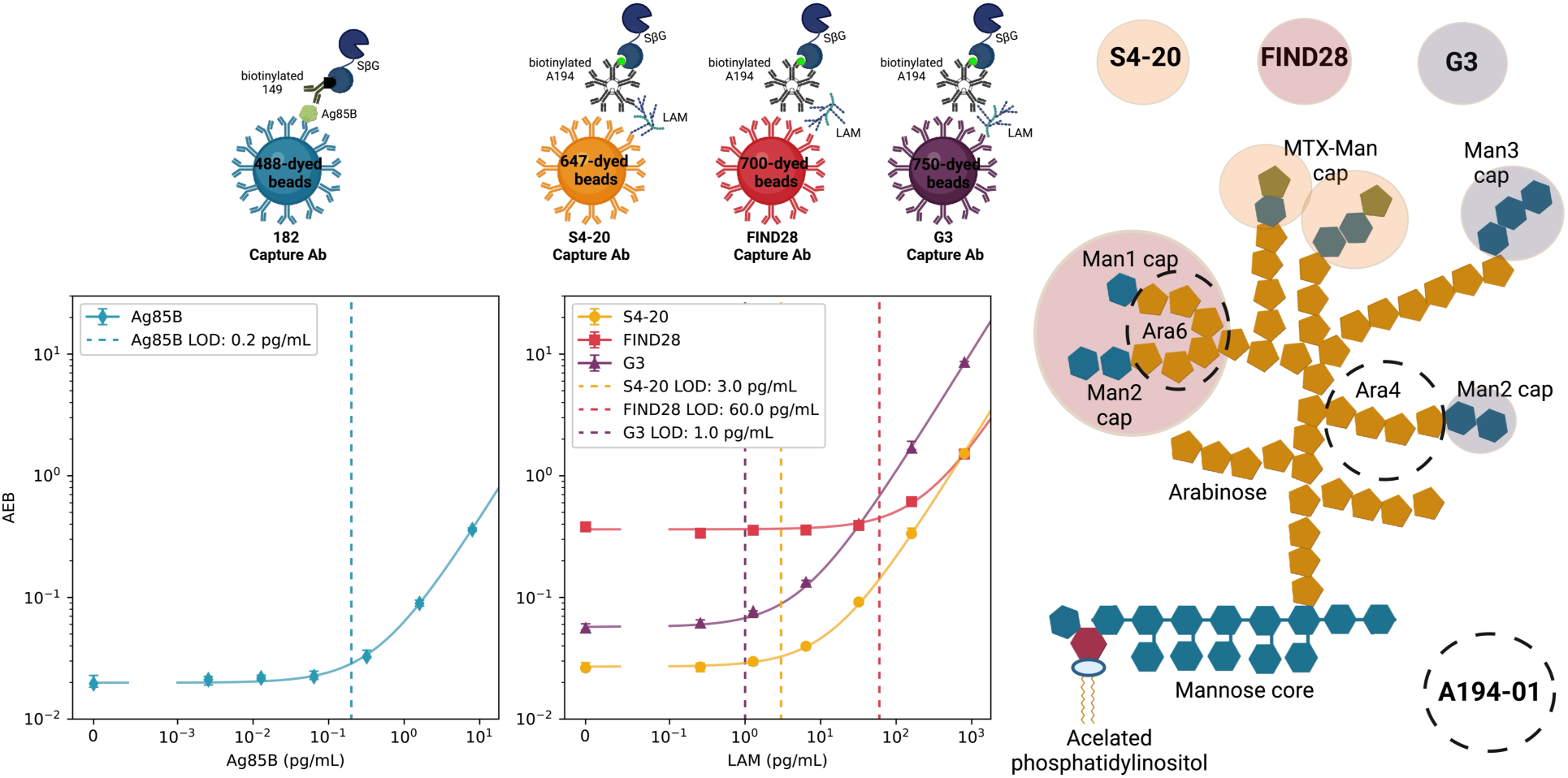
Multiplexed Simoa assays to quantify Ag85B and LAM in urine. For the Ag85B assay, 182λ coated 488 dye-encoded magnetic beads capture Ag85B in urine. For the LAM assays, S4-20, FIND28, and G3 antibodies are coated on 647, 700, and 750 dye-encoded beads and capture LAM in urine. Biotinylated 149 IgG antibodies and A194-01 IgM antibodies bind to Ag85B and LAM, forming sandwich assays. Streptavidin-β-galactosidase (sβG) binds the biotinylated antibodies and transforms its substrate, Resorufin-β-d-galactopyranoside (RGP), to its fluorescent form, which is detected by the HD-X instrument. A camera images and counts the number of fluorescent wells (“on” beads) (meaning they have a protein molecule bound) and the total number of wells containing a bead. The ratio of these values (average enzyme per bead; AEB) is converted back to a concentration according to the corresponding calibration curve.

The images were automatically analyzed by the HD-X image processing software to identify which wells contained beads, categorize beads by plex (fluorescent channel), and measure the intensity of each well in the resorufin channel. For each sample, it calculated fon, the fraction of beads with increased intensity in the resorufin channel; Ibead, the average resorufin intensity of “on” beads; and the average number of enzymes per bead (AEB) according to digital ELISA (using Poisson statistics) and analog ELISA (based on average intensity).

*Data processing*—Concentrations were calculated using calculated *f*_on_, *I*_bead_, digital AEB, and analog AEB in the run histories produced by the HD-X software. Analog AEBs for FIND28 (700 nm channel) were calculated according to Zhang et al. (2023), with an adjustment to the calculated mean fluorescence intensity of wells with single enzyme molecules, because the background fraction on (approximately 30%) was too high for the HD-X software to calculate it [33]. Replicate AEBs were calculated as a weighted average of digital and analog AEBs as in Zhang et al. (2023) [33]. Calibration curves were created by least-squares linear regression of AEB against the concentrations of known calibrators of Ag85B and LAM for each HD-X run. Using the appropriate calibration curves, each replicate of each sample was assigned an Ag85B concentration and three LAM concentrations, one for each capture antibody. The concentration for each sample in each plex was the median concentration of all replicates.

### 4. Assay development and validation

Simoa assays were developed and validated for 11 *M. tb* antigens in urine (Fig S3, Table S10). Two of these antigens (LAM and Ag85B) were detectable in most TB urine samples and undetectable in most non-TB urine samples (Table S4). These assays were then optimized, validated, and combined into a single multiplex Simoa assay.

To find the best antibody pairs for each target, a pairwise screening process was conducted using the HD-X analyzer and a pooled set of positive TB samples from the training cohort diluted with healthy urine. Available antibodies were paired as either capture reagents (conjugated to beads) or detection reagents (biotinylated). The antibodies were screened in a three-step format, and the SNR values were obtained. Optimal pairs showed a high signal-to-background ratio with a low background signal (Figure S2). A194 as a detector gave the highest SNR across most antibodies tested. A194-01 IgM (Rutgers University, Newark, NJ, USA) is best characterized and was shown previously to bind to urine LAM as a detector antibody (20, 22). Therefore, in addition to the antibody screening, other considerations when choosing the final antibodies included the reproducibility of the antibodies, the epitopes the antibodies bind to, and the isoelectric point (pI) of the antibodies. Considering all these parameters, the A194-01 was chosen as the detector for our LAM assays.

The capture antibodies chosen for LAM were G3, S4-20 (Otsuka Pharmaceutical Co., Tokyo, Japan), and FIND28 (FIND). The capture antibody chosen for Ag85B was the 182λ antibody, and the detector was the 149 antibody. The Ag85B antibodies were obtained from AbCellera Biologics (Vancouver, Canada) under material transfer agreements for Simoa assay development. Genscript was contracted to produce large batches of identified antibodies using sequence information provided by AbCellera Biologics under non-disclosure agreements.

A four-plex Simoa assay was developed and optimized by adjusting various assay parameters, including detection antibodies concentration, enzyme concentration, assay format, incubation times, and diluent buffers.

Assays were validated through dilution linearity; spike and recovery; and dropout tests. In dilution linearity, urine samples from three individuals with TB were successively diluted neat, 1:2, 1:4, and 1:8 in sample diluent to confirm that for each two-fold increase in dilution factor, the measured concentration decreased by a factor of two. In spike and recovery, urine samples from three individuals without TB were spiked with known concentrations of analyte standard, and the measured (recovered) concentration was compared to the known (spiked) concentration. In dropout tests, only one of the standards (LAM or Ag85B) was added each time to evaluate its effect on the measured concentration of the other biomarker.

### 5. Alere LAM assay

*Sample and Test Preparation*-Lateral flow test strips (Determine TB LAM Ag, Abbott 7D2741) were prepared by individually separating each test strip and removing the cover according to the manufacturer’s instructions. A total of 204 blinded human urine samples, previously stored at - 80°C, were thawed at room temperature. 80 µL of each sample was centrifuged at 2,000 × g for 10 minutes at 4°C. For each sample, 60 µL of the supernatant was pipetted from the tube and applied to the sample pad on the corresponding test strip. The test strips were allowed to sit in a dry, room temperature environment for 25 minutes before interpreting the results.

*Interpretation of Results-* Samples were classified as LAM negative if the “control” line on the test strip was present, but no visible line appeared in the “patient” section. Conversely, samples were considered LAM positive if both the “control” line and the “patient” line both were visible. In cases where the “control” line was present and the “patient” line was exceptionally faint, the results were noted but treated as positive cases.

### 6. Statistical analysis

*Machine learning*—Cross-validation within the model-building cohort (120 samples) was used to select the model and narrow hyperparameters. Receiver-operating-characteristic area under the curve (ROC-AUC) was used as the primary performance metric, but balanced accuracy, F-score, sensitivity at ≤1 false positive, and specificity at ≥90% sensitivity were also considered. Hyperparameters were tuned using repeated stratified 5-or 10-fold cross-validation; results under other numbers of folds, ranging from 2 to 55, were used for comparison. Based on performance in the model-building and validation cohorts, a generalized additive model (GAM) of splines was selected after comparing the performance of logistic regression, random forest, and gradient boosting classifiers. For the spline, concentrations were transformed according to *X*_t_ = 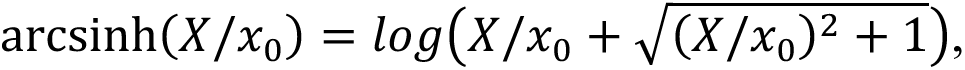 where x_0_ is 1/10 the limit of detection. This resulted in approximately normal features. Feature selection was performed by comparing performance using all possible subsets of the four biomarker levels; the use of all four biomarkers was chosen based on noninferiority, though in some cases in practice, the regularization led one or more features to have no influence.

*Blinded test set*—Hyperparameters (regularization strength, number of knots, and monotonic constraints for each feature) were chosen based on cross-validation within the training and validation cohorts. The final model is available online; see “Data and materials availability” below. The GAM returns a continuous score between 0 and 1, so based on cross-validation within the training and validation cohorts, scores above 0.72 were considered positive, indicating a TB diagnosis according to the model. The model and the corresponding predictions for the blinded test set were locked on November 4, 2024; on this date, the researchers submitted the Simoa concentration and predictions to representatives of the FIND specimen bank. The next day, November 5, 2024, FIND provided the researchers with the unblinded clinical results for this cohort, and the researchers proceeded to evaluate the model’s performance. No modifications, additions, or exclusion were made to the test data set from the point at which the model was locked down, and neither the test set nor any subset of it had ever been used to assess or refine the model being tested.

## RESULTS

### 1. A multiplexed assay for Ag85B and multiple forms of LAM in urine

We developed an ultrasensitive multiplexed Simoa assay to quantify Ag85B and LAM in urine. These analytes were chosen from a panel of 11 *M. tb* antigens for which Simoa assays were developed and tested in urine from individuals with and without TB (Tables S3 and S4, Figure S12). The assay format is depicted in Figure 1. Ag85B is measured with a pair of monoclonal antibodies at a limit of detection (LOD) of 0.2 pg/mL; LAM is captured by three different monoclonal antibodies that bind to distinctly different epitopes: S4-20 (MTX-Man caps), FIND28 (Ara6 with or without Man caps), and G3 (Man caps without MTX) at LODs of 3, 60, and 1 pg/mL respectively. A single detection antibody, A194-01 IgM, binds to Ara4 and Ara6 moieties with or without Man1 and MTX caps (Figure 1). These antibody pairs for LAM were chosen after extensive cross-testing of available antibodies using pooled urine from individuals with and without TB (Figure S13). They include the antibody pairs used by FujiLAM and MSD (S4-20/A194-01) and the Simoa LAM serum assay from Brock et al. (FIND28/A194-01) [20, 30], but the diagnostic measurement of different LAM channels plus Ag85B is unique to this work. Concentrations are determined from the fluorescent signal readout using linear calibration curves, so each sample is given one concentration for Ag85B and three concentrations for LAM, one for each capture antibody clone.

Urine samples are diluted twofold, so the analytical limits of detection of 0.2, 3, 60, and 1 pg/mL correspond to 0.4, 6, 120, and 2 pg/mL in urine samples. The assays do not cross-react: the presence of LAM does not affect the measured Ag85B concentration or vice versa (Figure S1, SI). Urine samples dilute linearly into the sample diluent (parallelism) and also when blended with other urine samples (admixture linearity), suggesting no differences in the detector antibody binding affinity to endogenous analytes and standard analytes and showing the flexibility of the assay at varying dilutions (Figures S2 and S3, SI). Specificity for native Ag85B was confirmed via immunoprecipitation and silver stain in wildtype and Ag85B deletion mutant BCG (Figure S4, SI). When known quantities of Ag85B and LAM were spiked into urine samples, the measured LAM concentrations were confirmed to equal the sum of the endogenous and spiked concentrations, with recoveries averaging 107% for S4-20, 103% for FIND28, and 130% for G3. The measured Ag85B concentrations were lower than the sum of the endogenous and spiked concentrations, with an average recovery of 54% (Figure S5, SI). The recovery of Ag85B was negatively affected by the urea concentration in the urine, especially at low-protein concentration samples (Figure S6, SI). We found that the urea in the urine attenuated the signal of the assay (Figure S7, SI). Ag85B renaturation attempts using TMAO did not yield higher recoveries. Adding urea to the calibrators improved the recoveries to some extent (Figure S8, SI). However, the urea concentration between individuals varies, with concentrations ranging between ∼17–166 mM, independent of Ag85B concentrations (Figure S7, SI). Urea was not added to the calibration curve because any one concentration would not translate well across different samples.

### 2. Accuracy in diagnosing TB

Using this multiplex assay for Ag85B and three different LAM forms, we measured samples from 576 individuals (Table 1). Each urine sample corresponded to a unique adult with symptoms suggestive of pulmonary TB. All urine samples were collected before treatment. All three cohorts were approximately evenly split between men and women and people with and without HIV. The training and validation cohorts were predominantly smear-positive, while the test cohort was exclusively smear-negative (Table 1). The test cohort was fully blinded: the experimenters had access to the numbers of positive and negative samples but not to any information about each sample until after the model was evaluated.

Proceeding in multiple stages (Figure S9, SI), we trained a generalized additive model (GAM) classifier to diagnose TB [34]. After model selection, a model was trained on the model-building and validation cohorts (361 samples) and evaluated on the blinded test cohort (215 samples) (Figure S9, SI; Figure 2, green line). Finally, repeated nested stratified fivefold cross-validation was used on the entire cohort (576 samples) to generate the most robust estimates and confidence intervals for accuracy metrics (Figure 2, black lines and gray ranges). In this nested cross-validation, 20% of the samples were set aside for testing; cross-validation was used on the remaining 80% to choose the model hyperparameters that best balanced bias and variance; a model was trained on the 80% with the best hyperparameters and tested on the remaining 20%. Repeating this process many times provided an unbiased estimate of the performance of our diagnostic test without data leakage, as well as minimally biased confidence intervals [35, 36]. The model performance characteristics in the nested cross-validation are shown in Table 2. The blinded ROC curve aligns with the nested CV curve for smear-negative patients (Figure 2).

**Figure 2.**
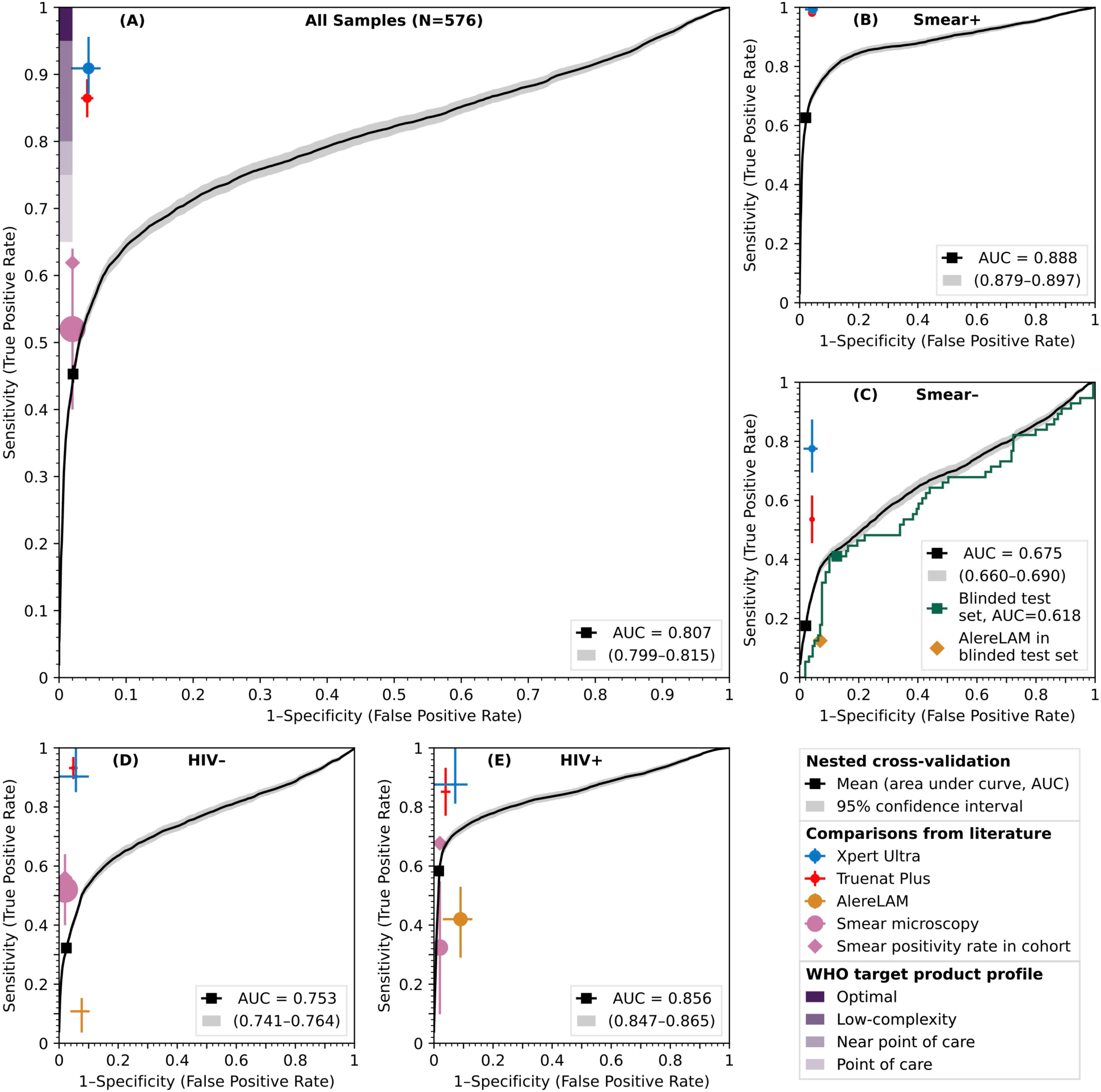
The model’s performance, stratified by HIV status and smear result. (A) **overall, (B)** smear-positive TB versus non-TB, **(C)** smear-negative TB versus non-TB, **(D)** people without HIV, **(E)** people with HIV. The black line and associated gray range give the mean and 95% confidence interval across all samples in the cohort, using nested cross-validation (CV). Squares give the empirical sensitivity and specificity at predetermined thresholds. The WHO optimal and minimal ranges are depicted in shaded purple rectangles in the upper right-hand corner. Xpert Ultra, Truenat Plus, AlereLAM, and smear microscopy performances and confidence intervals are depicted by blue, red, gold, and salmon crosses, respectively, with larger circles corresponding to larger cohorts. The actual rates of smear positivity in the cohorts are indicated with salmon diamonds. In panel **(C)**, the model’s performance in the blinded test set after training on the training and validation cohorts is shown in green, and performs similarly to the results from nested CV. The green square indicates sensitivity and specificity at predetermined thresholds, and the gold diamond gives the empirical performance of AlereLAM in the blinded test set.

**Table 2.**
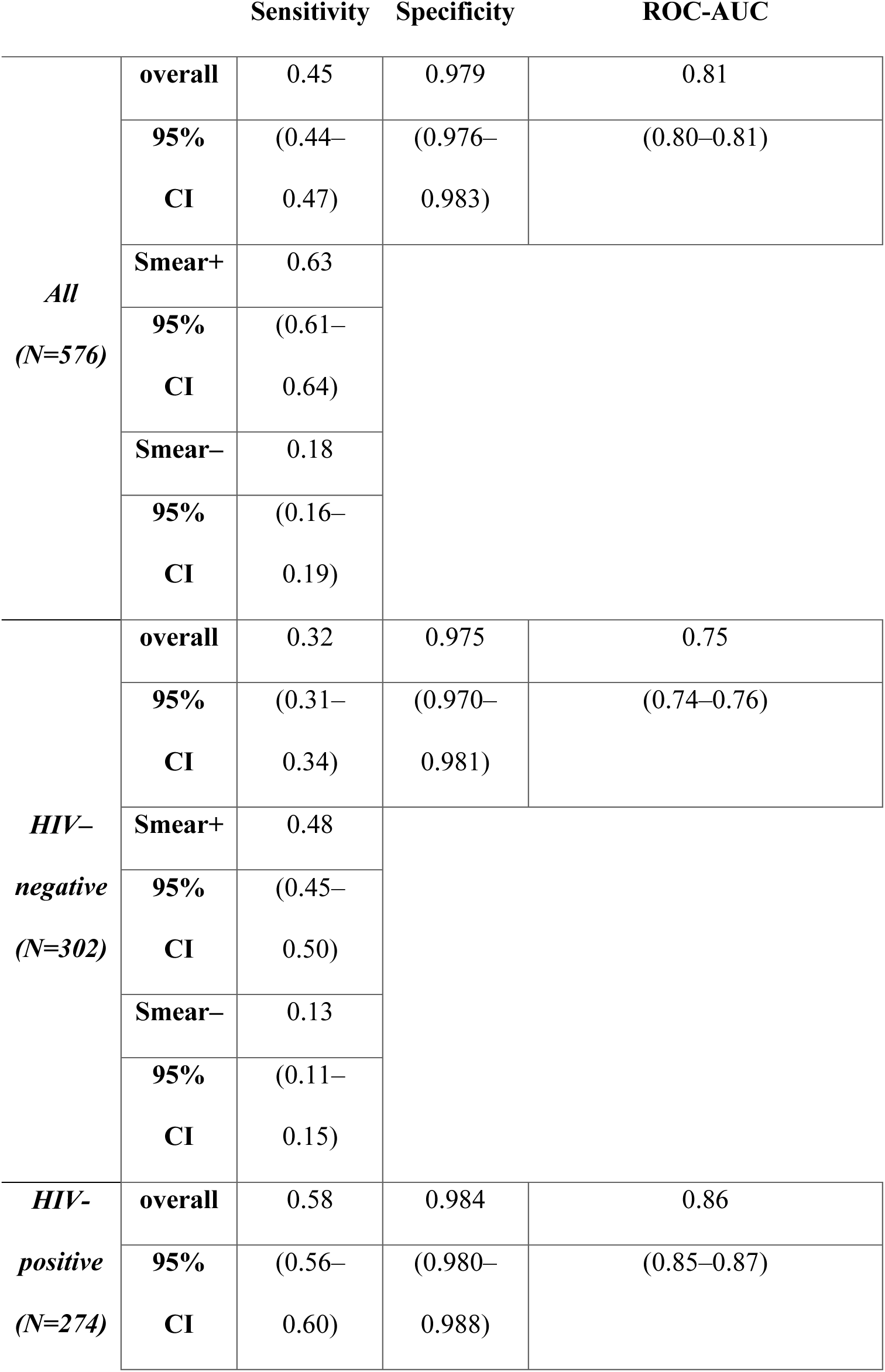
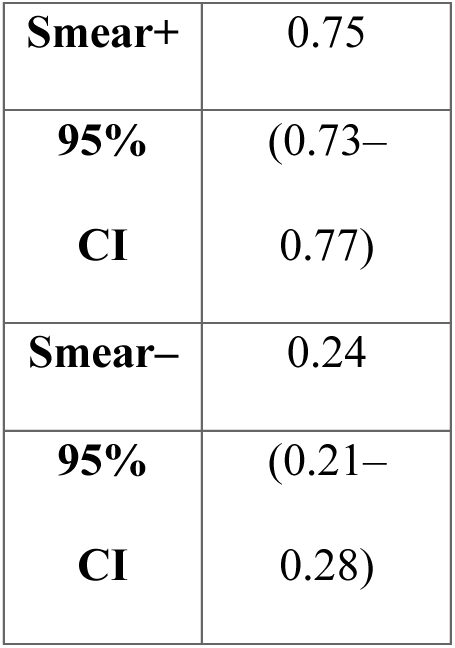
Model performance characteristics in nested cross-validation.

|  |  | Sensitivity | Specificity | ROC-AUC |
| --- | --- | --- | --- | --- |
| *All*<br>(N=576) | **overall** | 0.45 | 0.979 | 0.81 |
|  | **95% CI** | (0.44–0.47) | (0.976–0.983) | (0.80–0.81) |
|  | **Smear+** | 0.63 |  |  |
|  | **95% CI** | (0.61–0.64) |  |  |
|  | **Smear–** | 0.18 |  |  |
|  | **95% CI** | (0.16–0.19) |  |  |
| *HIV–*<br>*negative*<br>(N=302) | **overall** | 0.32 | 0.975 | 0.75 |
|  | **95% CI** | (0.31–0.34) | (0.970–0.981) | (0.74–0.76) |
|  | **Smear+** | 0.48 |  |  |
|  | **95% CI** | (0.45–0.50) |  |  |
|  | **Smear–** | 0.13 |  |  |
|  | **95% CI** | (0.11–0.15) |  |  |
| *HIV–*<br>*positive*<br>(N=274) | **overall** | 0.58 | 0.984 | 0.86 |
|  | **95% CI** | (0.56–0.60) | (0.980–0.988) | (0.85–0.87) |

|  |  |
| --- | --- |
| **Smear+** | 0.75 |
| **95% CI** | (0.73–0.77) |
| **Smear–** | 0.24 |
| **95% CI** | (0.21–0.28) |

In Figure 2, the model’s performance in various subsets is shown in receiver operating characteristic (ROC) plots, showing true positive rate (sensitivity) versus false positive rate (1-specificity). Overall, the sensitivity of our assay is 45% (95% CI: 44%–47%) with 98% specificity (95% CI: 97.6%–98.3%) (Table 2). The contribution of having a four-plex assay versus other combinations was also evaluated by calculating the AUC-ROC scores for each individual biomarker (Figure S11, SI). Adding Ag85B to all other combinations improved the AUC-ROC scores (Figure S11, SI). Including HIV status, sex, and age as predictors did not improve the performance of the model in initial tests, so they were not used. According to our estimated TB probability, nine out of 17 samples classified as Clinical TB (∼50%) were detected as positive TB, and 26 out of the 27 samples classified as “Likely subclinical (96%) TB” were detected as negative TB.

### 3. Ag85B and LAM concentrations in urine

Figure 3 shows swarm plots for the concentrations of Ag85B and LAM (according to the three capture antibodies). The concentrations of all biomarkers are higher in the TB samples than in the non-TB samples. The LAM concentration differs for each capture antibody, as it identifies different epitopes on LAM, and their prevalence differs.

**Figure 3.**
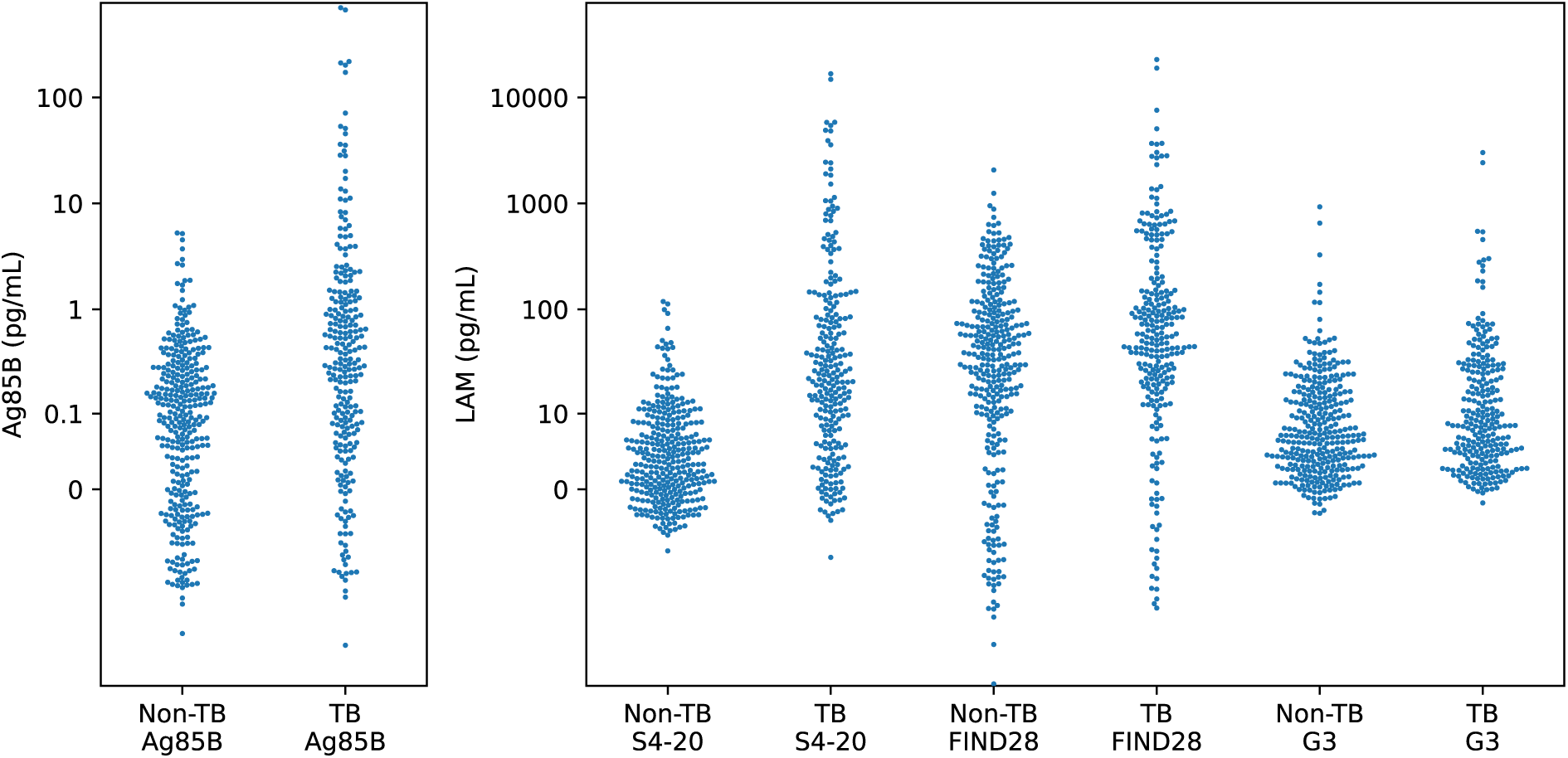
Ag85B and LAM concentrations in urine. Each sample is represented by a dot. For each of the four assays (Ag85B and three LAM capture antibodies), results are separated by TB diagnosis.

The LAM concentration according to S4-20 contributes the most to the model’s decision (Figure S11). The addition of Ag85B slightly improves the model, and G3 and FIND28 contribute less (Figure S11).

Significantly higher concentrations of all biomarkers were detected in HIV+C+ samples from Vietnam compared to South Africa and Peru. For HIV-C+ samples, significantly higher levels of LAM detected by FIND28 were measured in South Africa and Peru compared to Cambodia. (Figure 4).

**Figure 4.**
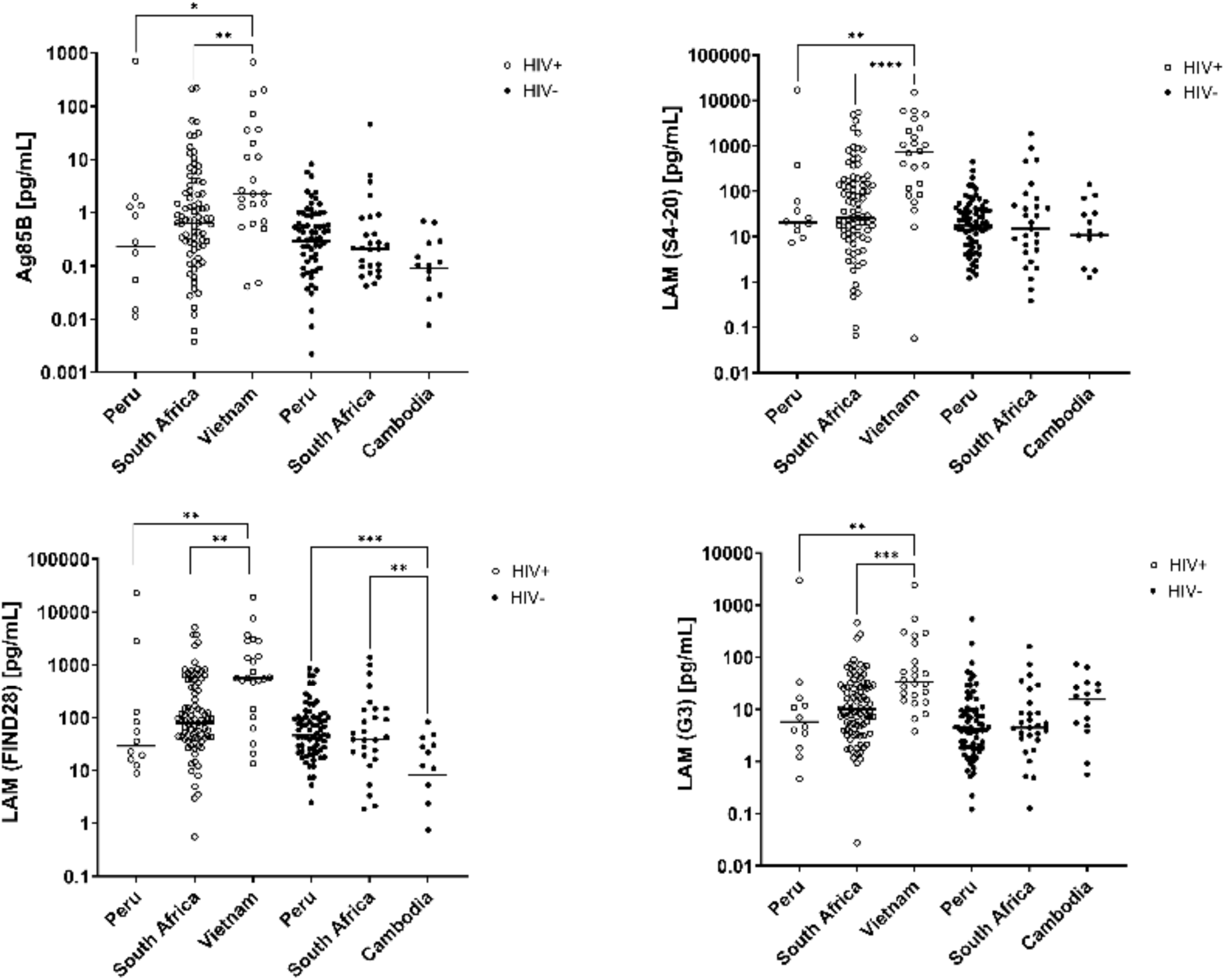
Ag85B and LAM concentrations in urine stratified by country and HIV status. The concentrations of Ag85B and LAM detected by three capture antibodies (S4-20, FIND28, and G3) are plotted for each country of origin (Peru, South Africa, Vietnam, and Cambodia) according to HIV status. All samples are classified as either culture-positive or clinical TB. The black line is the median. Statistical analysis was done using One-Way ANOVA with the Kruskal-Wallis non-parametric test. One asterisk represents p≤0.05. Two asterisks represent p≤0.01. Three asterisks represent p≤0.001. Four asterisks represent p<0.0001.

The results of Alere LAM using 215 samples from the blinded test cohort were compared with the results of our multiplex assay (Table 3). Overall, the sensitivities of AlereLAM and the multiplex Simoa assay were 12.5% (7/56) and 41% (23/56), and the specificities were 93% (148/159) and 87.4% (139/159). Similar results for the Alere test were seen using 244 samples (Table S2). Because AlereLAM is approved for HIV-positive patients only, we also stratified the samples by HIV status. For HIV-positive patients, the sensitivities of AlereLAM and the multiplex Simoa assay were 23.8% (5/21) and 28.6% (6/21), and the specificities were 93.9% (62/66) and 95.5% (63/66).

**Table 3.**
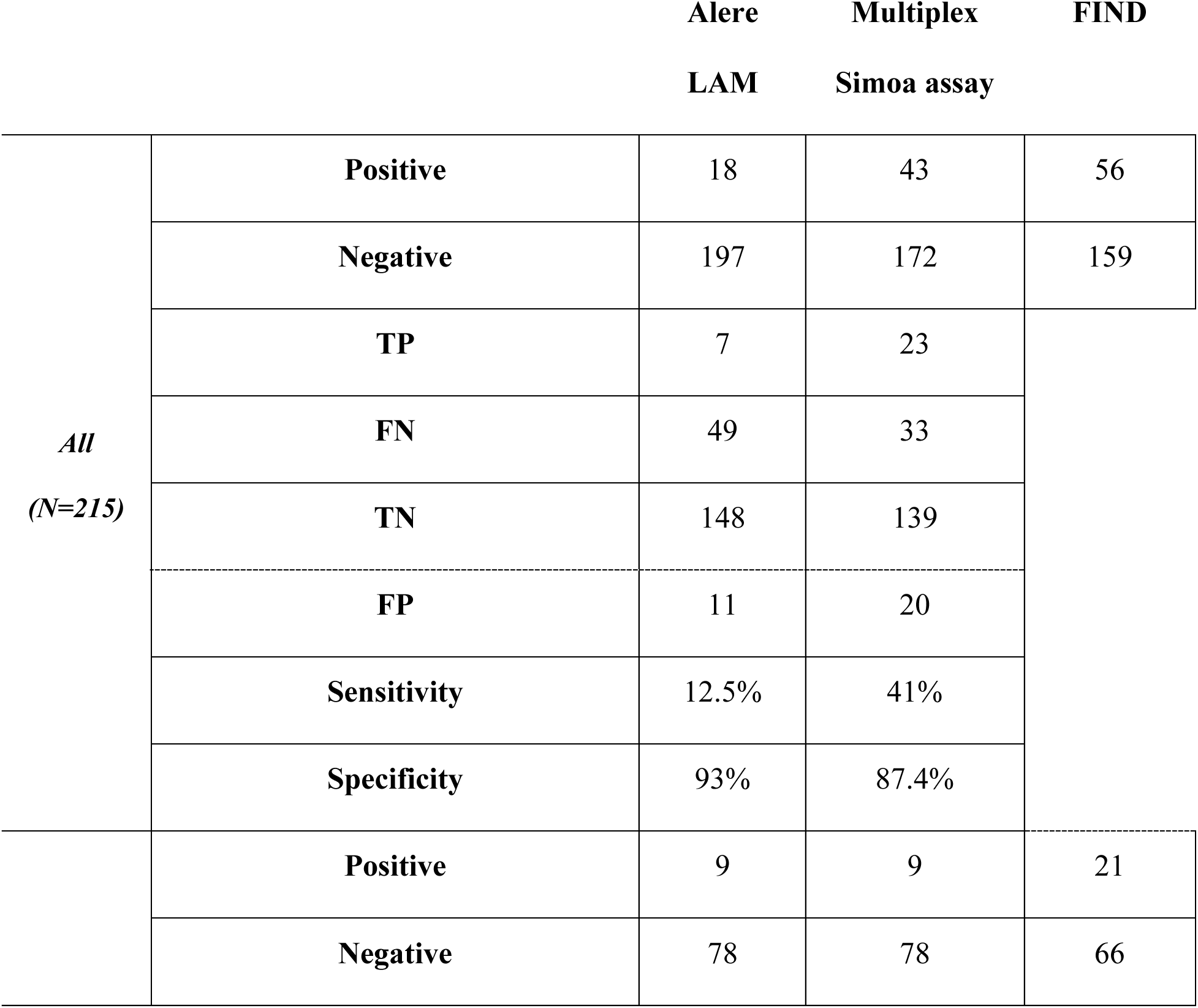
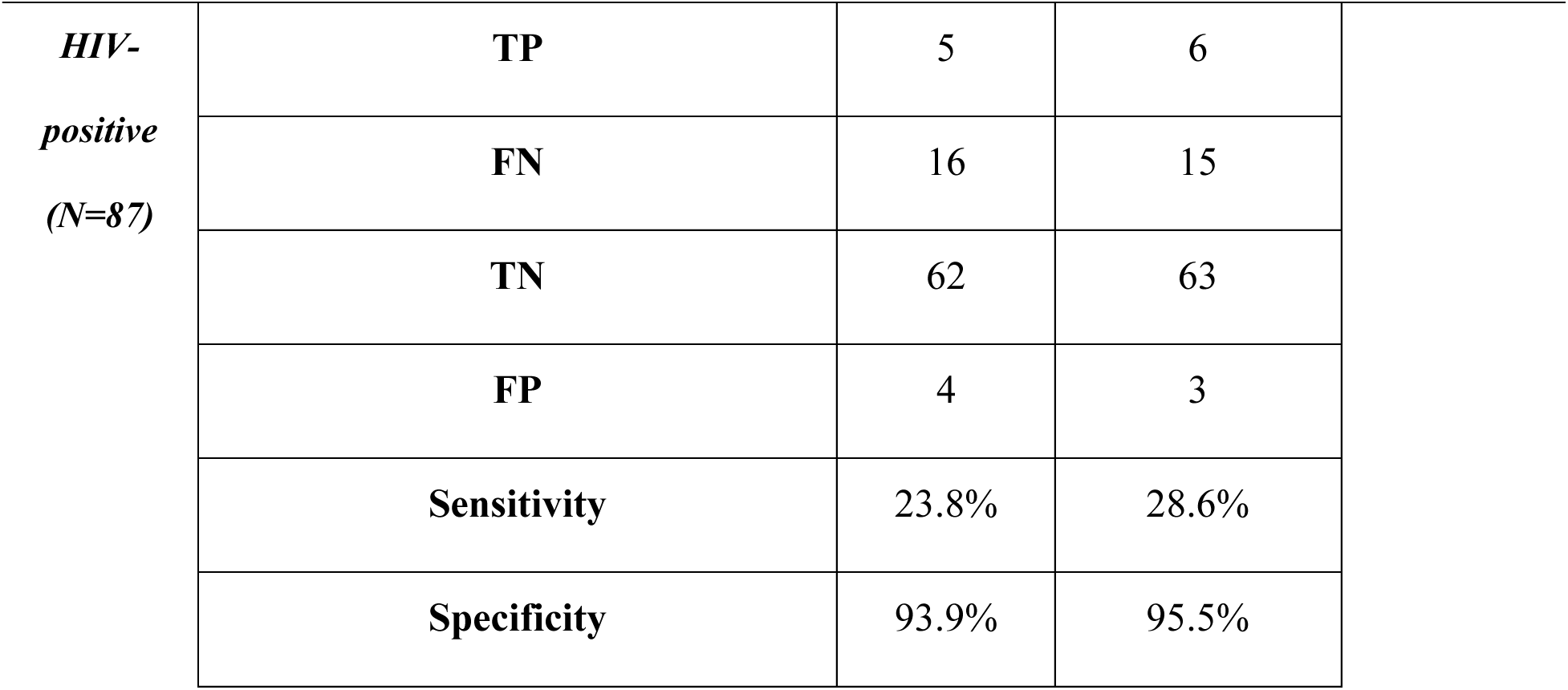
Comparison of Alere LAM results and Simoa multiplex assay.

## DISCUSSION

Although TB is curable using antibiotics, it is still the leading infectious cause of death worldwide. The main challenges related to the diagnosis of TB include the use of biohazardous sputum, insufficient sensitivity, and slow turnaround time for the gold standard culture. Here, we present an ultrasensitive multiplex biomarker assay in urine that could be used as an adjunctive test to diagnose TB.

Our test includes the protein biomarker Ag85B and three capture antibodies for the glycolipid biomarker LAM. As these three capture antibodies identify different epitopes of LAM, we hypothesized that combining the results of all three binding pairs would improve the assay’s performance. Concentrations for all three capture antibodies are calculated using the same LAM standard, but they perform quite differently: S4-20 best separates people with and without TB, FIND28 measures the highest concentrations in urine on average, and G3 has the best analytical sensitivity. S4-20 was less susceptible to both background nonspecific binding (Figure 1) and cross-reactivity with components present in the urine of people without TB (Figure 3), so it is understandable that it contributed most to the model.

The 2024 edition of the TPP did not relax the 2014 edition’s criteria for sensitivity or specificity [17, 18]. But in the intervening decade, new diagnostics have been introduced that fall below its stringent minimum specificity (98%) relative to culture: Xpert Ultra (96%), AlereLAM (91%), and Truenat Plus (96%) [2, 14–16]. This reflects the importance and difficulty of detecting TB, as well as the limitations of culture as a gold standard: some “false positives” according to index assays may well be false negatives of the reference assay [37]. However, although our test is less sensitive than Xpert Ultra and Truenat Plus, it uses safe and accessible urine samples, rather than sputum. Our assay is a first step towards a diagnostic test that may supplement existing diagnostics and aid clinicians in deciding whether to start TB treatment. For example, the AlereLAM assay is recommended for HIV-positive patients, although its sensitivity is quite low (42% sensitivity at 91% specificity) [2]. Because our test has ∼60% sensitivity at 98% specificity among people with HIV, it can serve as a better diagnostic test for this population. On the other hand, because our assay has a sensitivity of 63% among smear-positive individuals at a specificity of 98%, it is not ready to replace smear microscopy. Additional aspects presented in the TPP include the test cost (which should be <US$ 8 per measurement for a low-complexity test), the test’s simplicity (three manual steps), and the assay’s duration (one hour) [18]. The current Simoa-based assay costs less than US$ 6 for a single replicate (but more for the three replicates used in this study), requires several manual steps and precise measuring (which could be streamlined during scale-up), and returns results in about two hours, similar to the Xpert and Truenat tests. The capital cost and instrumentation size could be reduced by porting the assay to a benchtop Simoa system like the Quanterix SR-X, or by converting the assay to MOSAIC, which uses a flow cytometer readout [38, 39]. The assay could be adapted to a readout device with fewer multiplex channels by narrowing the number of biomarkers and affinity agents (Figure S11).

If this study is generalizable, our assay has diagnostic accuracy (sensitivity/specificity) modestly superior to AlereLAM; inferior to culture, Xpert Ultra, and Truenat Plus; and overlapping with smear microscopy (Figure 2). It would be comparable to an updated FujiLAM assay (2024, called FujiLAM II or FujiLAM2) which purports to solve the aforementioned lot-to-lot variability; a retrospective study demonstrated 80% sensitivity (55/69 samples) and 93% specificity (80/86 samples) in people with HIV, while a prospective preprint reported lower sensitivity in TB meningitis [40, 41]. In addition, it is more sensitive but slightly less specific than the serum LAM Simoa assay developed by Brock et al [30].; less sensitive in HIV-negative people than the MSD EclLAM assay developed by Sigal et al [42]; and more specific than the Simoa cytokine triage test developed by Ahmad et al. [31](Neither the new FujiLAM studies nor Ahmad et al. reported smear positivity rates, which are a major determinant of ease of diagnosis.)

This study has several limitations. It employed a retrospective case-control study design, preventing determination of positive and negative predictive values in a real-life context. Such a design also risks overestimating performance due to the potentially biased inclusion of patients at extremes of phenotype distribution that might not represent the target population (spectrum bias) [37]. We attempted to mitigate this by including individuals classified as “likely subclinical TB” and “clinically diagnosed with TB.” Samples were chosen based on target numbers of HIV, culture, smear, and latent TB status, introducing a selection bias. Despite efforts to stratify, our cohort may not be representative of all people who present for TB testing at a given center, or of the combined populations in the meta-analyses for other tests. Because our cohort was chosen to include a certain percentage of smear-positive and smear-negative samples, a direct comparison with the literature-reported performance for smear microscopy is problematic. (Because AlereLAM in the blinded test cohort was conducted alongside but independently of Simoa, it is a more appropriate comparator, subject only to the biases inherent in the study design.) Case-control diagnostic studies are considered less generalizable than prospective studies, but as long as data collection is planned before testing and samples represent the full spectrum of the disease, they have not been found to have a consistent bias relative to prospective studies [43].

Another limitation is the age of the samples used. Out of 576 samples, 77 were frozen from 2012–2015, and 511 were frozen from 2016–2019. Using fresh samples or samples frozen for a shorter period in a prospective study might improve assay performance.

We performed a blinded validation to evaluate the model’s performance. However, the TB samples from the test cohort were all smear-negative, so we did not have a truly blinded validation of our assay’s performance in the roughly half of people with TB who are smear-positive. The strong correspondence between results in the blinded test cohort and nested cross-validation (green and black lines, respectively, in Figure 2) is consistent with theoretical and simulated findings that nested cross-validation is an unbiased estimator of model performance [36]. Further independent validation in a prospective, real-world setting is warranted to assess the true performance of this diagnostic test.

Significant heterogeneity was observed in biomarker concentrations, some of which was explainable by country (Figure 4). HIV-negative patients with a positive culture test from Cambodia had lower levels of LAM according to the FIND28 assay, compared to South Africa and Peru. The lower levels measured in Cambodia may be attributed to the different lineages in each country. *M.tb* has seven lineages. In South Africa and Peru, the dominant lineage is lineage 4, in Vietnam lineages 1 and 2 [44], and in Cambodia lineage 1 [45]. The differences in the dominant epitopes present in each country may imply that different LAM forms are present in the urine of patients from different countries with different lineages. It is possible that lineages 4 and 2 (South Africa, Peru, and Vietnam) have predominantly LAM molecules with Ara6 cap with or without any Man cap (FIND28). Other reports show that there are different ratios of the LAM epitopes in urine [21]. For example, urine samples from Peru and South Africa showed a 4-fold higher LAM amount using FIND28 versus S4-20 antibodies as capture antibodies [46], which may be due to the prevalence of specific epitopes in LAM to which each antibody binds, and the heterogeneous nature of LAM structures in urine [46]. In future studies, such as the prospective validation of the present assay, it would be instructive to conduct mycobacterial sequencing and compare biomarker abundance results with lineage and phenotype information.

One of the major hurdles for controlling TB is the patient’s delay in seeking treatment. For example, a four-week delay in seeking treatment was seen in KwaZulu Natal, South Africa, and a ten-week delay in South Africa’s rural Northern Province, now officially known as Limpopo, which far exceeds the WHO-recommended two weeks for initiating treatment after suspicion [47]. Reasons for delay include distance from diagnostic facilities, long waiting times, and absence of clinical symptoms [47, 48]. Delays in diagnostic testing in the different countries can be part of the cause for lower concentrations of biomarkers in the urine at the time of sampling. It may be valuable to record the number of days from the onset of symptoms to seeking a diagnosis, especially when acquiring samples for clinical trials, as the concentration of biomarkers can be affected. (FIND collected a “binned” version of symptom duration for some individuals in our study, but the data were too coarse for this type of analysis.) In addition, this information may aid in assessing pre-test probabilities.

In 2022, FIND reclassified TB into the following stricter categories: TB; S+C+, TB; S-C+ TB; S-C-(CXR) TB; S-C-(Molecular/Xpert); and Non-TB. Another classification in our cohort was “likely subclinical TB”, which refers to patients with positive chest X-rays at enrollment. These patients did not begin treatment at enrollment, but they had worsened symptoms and a positive culture at follow-up. “Clinical TB” refers to patients treated based on their symptoms who did not have a positive diagnostic result, no TB history, negative smear and culture results, and a negative QuantiFERON-TB Gold (QFT) result used to assess LTBI. Our test successfully identified TB in half of the clinical TB samples, suggesting that it would help diagnose cases missed by other tests. Clinicians could use our test to make treatment decisions when all other diagnostic tests, such as AlereLAM, give negative results. In addition, the ability to detect 96% of the “likely subclinical” samples as negatives may suggest that our assay can be used in the future as a confirmatory test for this population. Of course, a larger cohort of this population needs to be validated first.

## CONCLUSIONS

We have demonstrated that the measured LAM concentrations in urine depend on the antibodies used to measure them. By employing multiple antibody pairs, we developed a multiplexed assay for LAM and Ag85B that is more accurate because it accounts for these different binding specificities. In many people with TB, LAM is below the detection limit of an ultrasensitive assay, suggesting that increasing analytical sensitivity is not sufficient to achieve further clinical sensitivity. We hope that the present test can be translated into a clinical laboratory assay using MOSAIC to conduct readout with a flow cytometer, and that its large set of LAM and Ag85B measurements can serve as the groundwork for the development of other diagnostics for TB.

### List of Supplementary Materials

Ag85B pull-down and silver staining

**Figure S1.**No cross-reactivity between the assays in dropout experiments.

**Figure S2.**Dilution linearity.

**Figure S3.**Admixture linearity.

**Figure S4.**Ag85B capture antibody (182λ) identifies the native form of Ag85B.

**Figure S5.**Spike and recovery of LAM and Ag85B.

**Figure S6.**Recovery of Ag85B is negatively affected by urea in the urine and is dependent on urine total protein.

**Figure S7.**Urea attenuates Ag85B concentration and spans a wide range in urine samples.

**Figure S8.**Attempts to improve recovery. Adding urea to the spiked matrix slightly improved the recovery of Ag85B.

**Figure S9.**Flow diagram of model training and evaluation.

**Figure S10.**Partial dependence plots for the model evaluated on the test cohort.

**Figure S11.**AUC-ROC scores with different combinations of biomarkers.

**Figure S12.**Calibration curves of all investigated markers.

**Figure S13.**Cross-testing LAM antibodies.

**Table S1.**Characteristics of common TB diagnostic tests.

**Table S2.** Alere LAM results using 244 samples from the blinded test set.

**Table S3.** TB biomarker Simoa assay characteristics. LOD: limit of detection.

**Table S4.** Detectability of 11 TB markers in the discovery cohort.

## DECLARATIONS

### Ethics approval and consent to participate

This study was exempted by Partners Human Research Committee (Protocol #2017P001447). All urine samples were provided by the FIND TB sample repository. FIND collected these samples under institutional review board (IRB)/independent ethics committee (IEC) approved studies in participating countries.

### Consent for publication

Not applicable

### Availability of data and materials

All raw data, as well as the code used to conduct the analysis and generate the figures and tables, will be available on GitHub.

### Authors’ contributions

Conceptualization: DRW, LX

Methodology: LX, TJD, SR

Software: TJD

Validation: TJD, SR

Formal analysis: TJD, SR

Investigation: TJD, SR, SCD, LX

Resources: DRW, TJD

Data curation: TJD

Writing—original draft: TJD, SR, LX

Writing—review & editing: DRW, SR, TJD, SCD

Visualization: TJD, SR

Supervision: DRW

Project administration: DRW

Funding acquisition: DRW, LX

## Data Availability

https://github.com/tylerdougan/simoa-tb

## Acknowledgments

We thank Bruce P. Bausk and Rabsa Sikder for conducting experiments that led to this study, as well as Ming Yang Lu and Faisal Mahmood of Brigham and Women’s Hospital for data analysis of the same. We thank Emmanuel Moreau of FIND for organizing and providing urine samples, Abraham Pinter of Rutgers University, and Masanori Kawasaki and Ryo Higashiyama of Otsuka Pharmaceutical Co. for kindly providing antibodies to LAM. We are grateful to Joel Ernst of UCSF for providing BCG cell lines, and Bryan Bryson of MIT for lysing them. We thank Olga Demler and Sebastian Cajas for their helpful suggestions on machine learning. Finally, we are grateful to the study participants who graciously offered biological samples and access to aspects of their clinical records.

Figure 1 was created in https://BioRender.com.

