## Supplementary Information for "A single-molecule array urine test for tuberculosis: A case-control diagnostic accuracy study"

### Development and validation of a multiplexed single-molecule array urine test for tuberculosis: A case-control diagnostic accuracy study

**Authors:**

Tyler J. Dougan[†1,2,3,4,5], Shira Roth[†1,2,3], Liangxia Xie[1,2,3], Sydney D'Amaddio[1,2,3], David R. Walt[*1,2,3]

**Affiliations:**

[1] Wyss Institute for Biologically Inspired Engineering, Harvard University; Boston, MA 02115, USA.

[2] Department of Pathology, Brigham and Women's Hospital; Boston, MA 02115, USA.

[3] Harvard Medical School, Harvard University; Boston, MA 02115, USA.

[4] Harvard-MIT Program in Health Sciences and Technology, Massachusetts Institute of Technology; Cambridge, MA 02139, USA.

[5] Present address: Africa Health Research Institute, Durban, South Africa.

† These authors contributed equally to this work

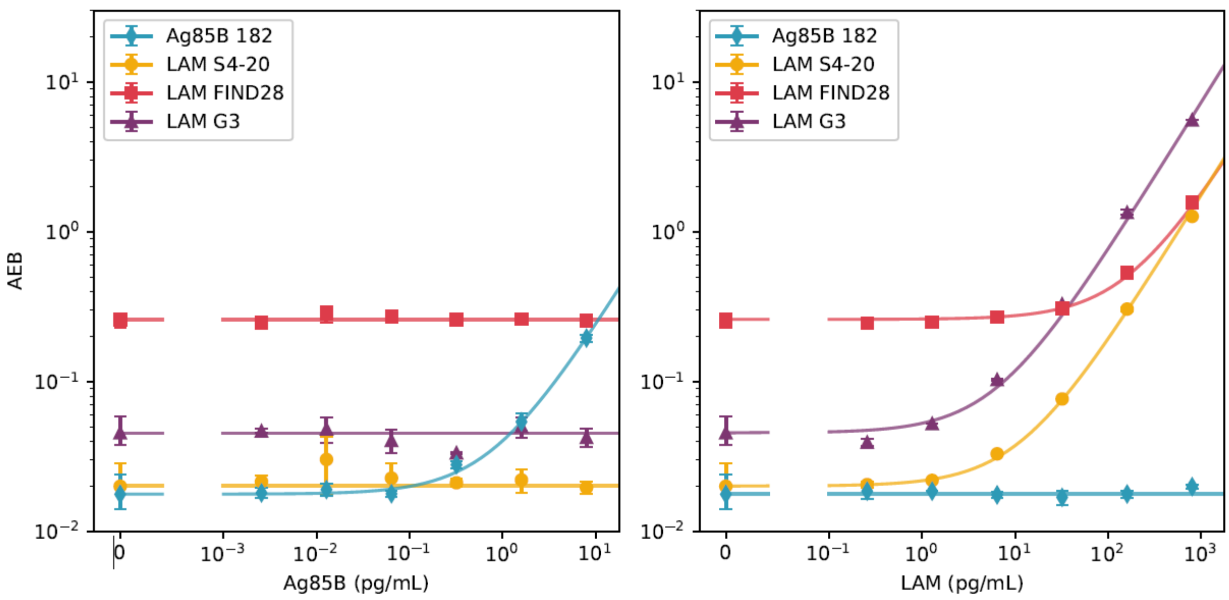


**Figure S1.** No cross-reactivity between the assays in dropout experiments. (Left) The presence of Ag85B did not affect the measured concentration of LAM. (Right) The presence of LAM did not affect the measured concentration of Ag85B.

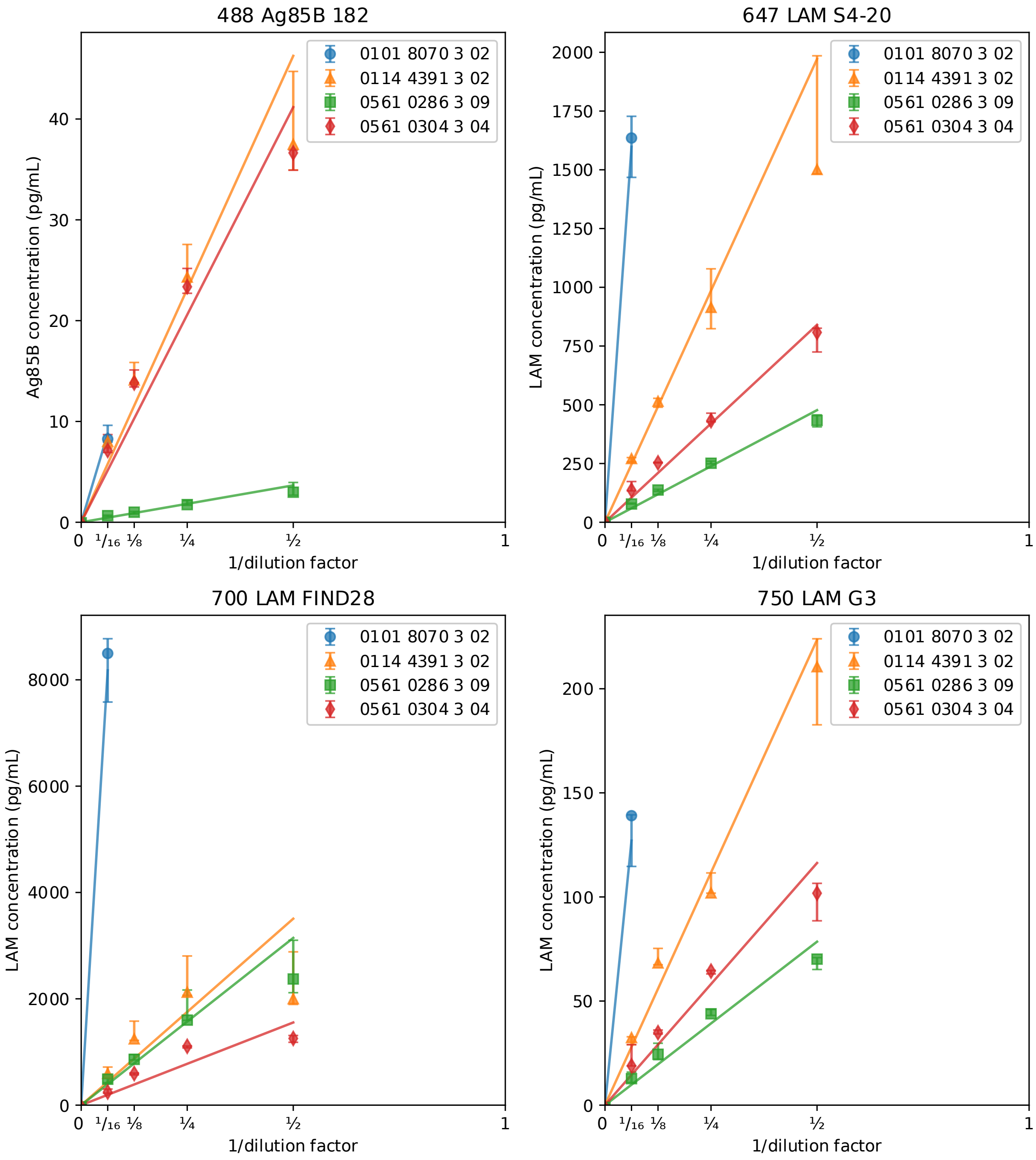


**Figure S2.**Dilution linearity. Urine samples (samples barcodes appear in the legend) were diluted 2X, 4X, 8X, and 16X with sample diluent.

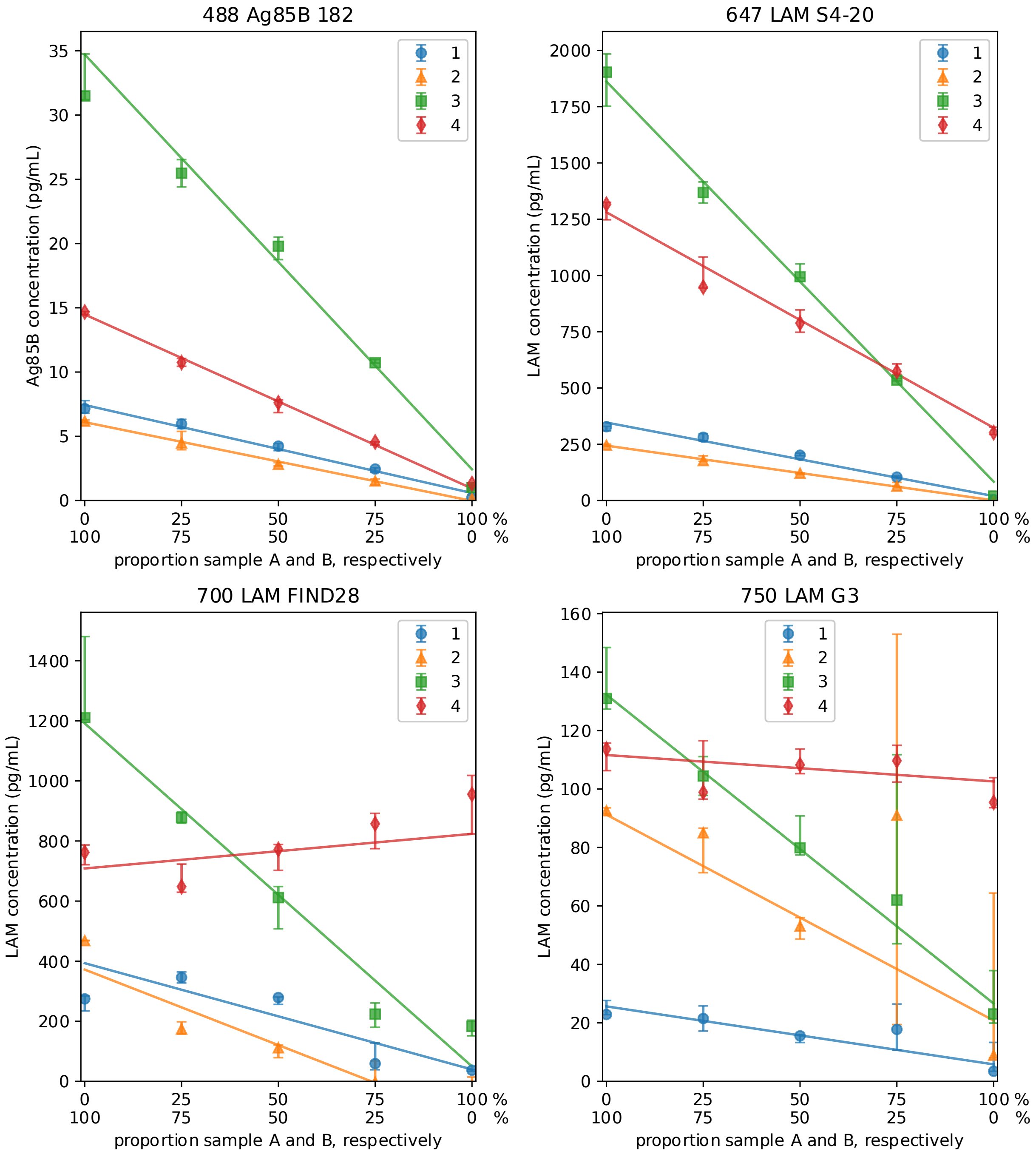


**Figure S3.** Admixture linearity. Urine samples were mixed according to the percentages indicated on the X-axis. Sample barcodes for the mixtures are as follows: 1- Sample A (01014262301), Sample B (05010163303); 2- Sample A (01610257301), Sample B(01610436301); 3- Sample A (01610485301), Sample B(01144309303); 4- Sample A (01610462301), Sample B(01610755301).

**Ag85B pull-down and silver staining**

To verify that the Ag85B capture antibody (182λ) used in this study identifies the native form of Ag85B, we conducted a pull-down assay using cell lysate from the BCG strain. It should be noted that there is a single amino acid substitution in the BCG copy of *fbpB* (expressing Ag85B protein) compared with the reference H37Rv sequence. A volume of 250 μL of each sample or RIPA buffer was mixed with 40 million 488-dyed magnetic beads conjugated with the anti-Ag85B antibody (182λ). Following an overnight incubation with rotation at 4°C, the samples were washed twice with RIPA buffer. Then, the beads were mixed with 3 μL of 500 mM DTT, 7.5 μL of 4X Laemmli buffer, and 19.5 μL of RIPA buffer. Following incubation of the beads at 95°C for five minutes, supernatants were loaded on a 4–12% precast gel (Novex™ WedgeWell™ 4–12% Tris-Glycine Gel) and run for one hour at 150V. The gel was silver stained according to the manufacturer's protocol (Pierce Silver Stain Kit, 24612). The Developer working solution was incubated for one minute to achieve the desirable protein band intensity. The gel was imaged using BioRad Gel Doc™ EZ imager.

Bands corresponding to Ag85B molecular weight (34.58KDa) are seen only in the positive control lane and the wild-type BCG cell lysate (marked in red rectangle), meaning that antibody-coated magnetic beads pulled down Ag85B from these lysates. These results show that the antibody identifies the native form of Ag85B. There is no band in the negative controls: Ag85B knockout, Ag85C, and without cell lysate. To differentiate the antibody light chain from Ag85B, the 182λ antibody was also run on the gel.

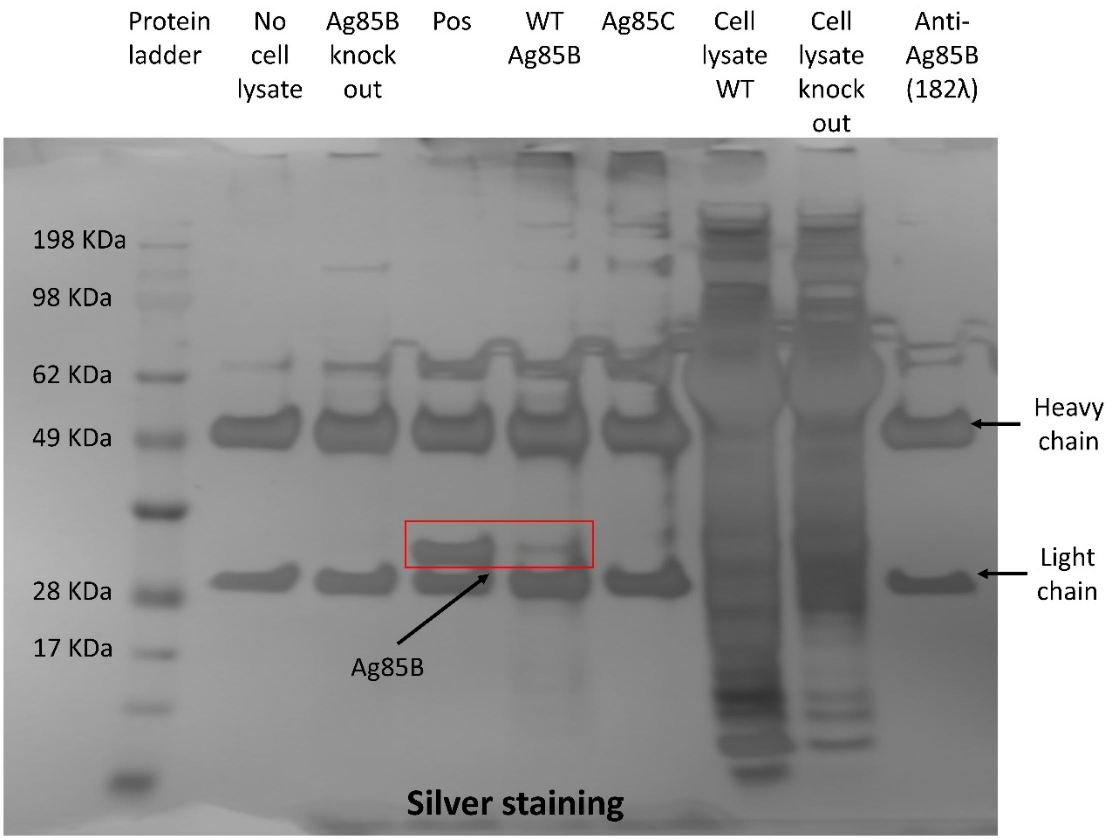


**Figure S4.** Ag85B capture antibody (182λ) identifies the native form of Ag85B. Silver stain of proteins pulled down with Ag85B capture antibody (182λ). Abbreviations: Pos, native Ag85B BEI; WT Ag85B, wild-type BCG cell lysate.

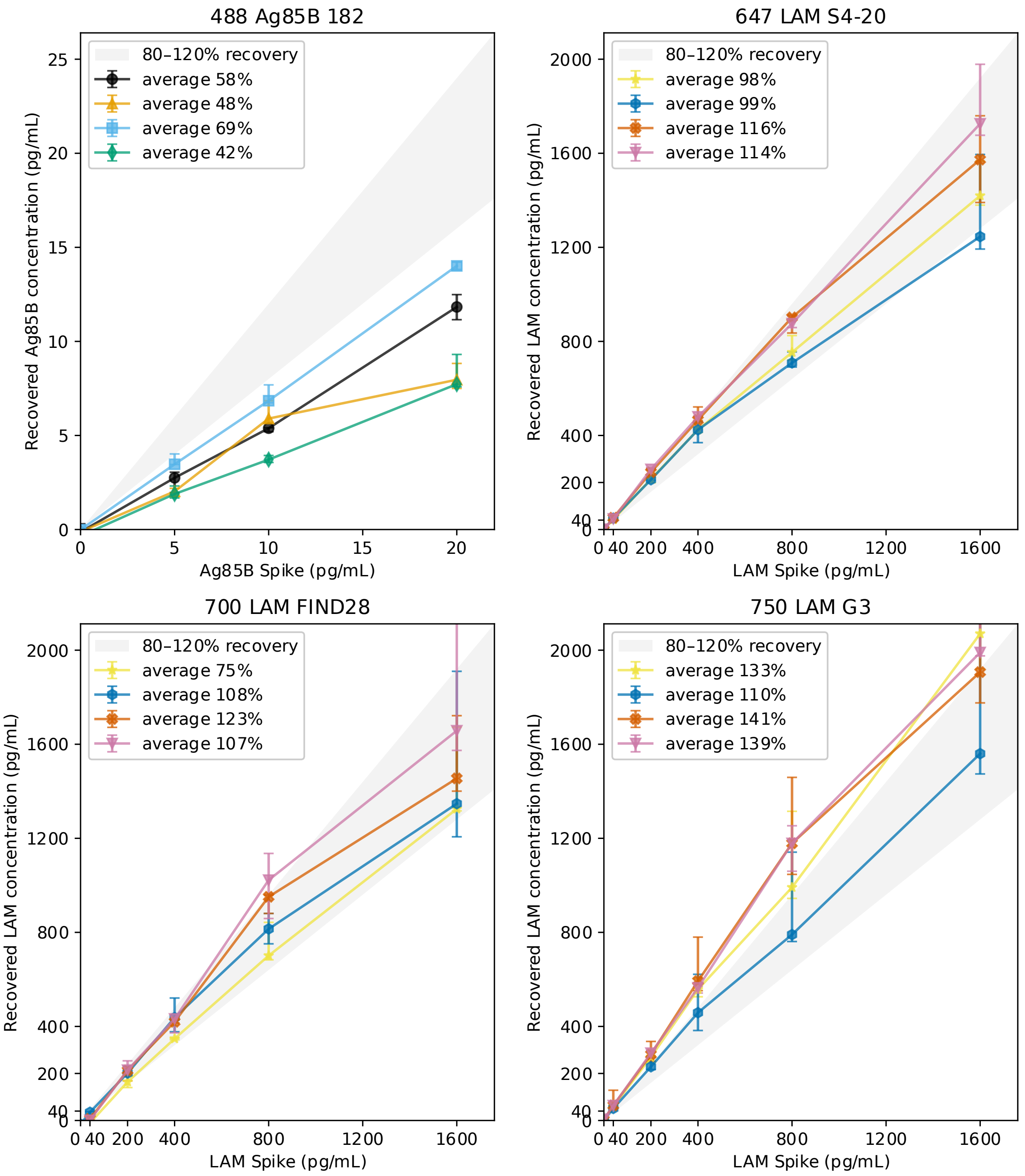


**Figure S5.** Spike and recovery of LAM and Ag85B. Known concentrations of Ag85B and LAM were spiked into negative urine samples from the training cohort. Recoveries were calculated as follows: $\% \, Recovery = 100\% \times \frac{Observed\ concentration - Endogenous\ concentration}{Spiked\ concentration}$.

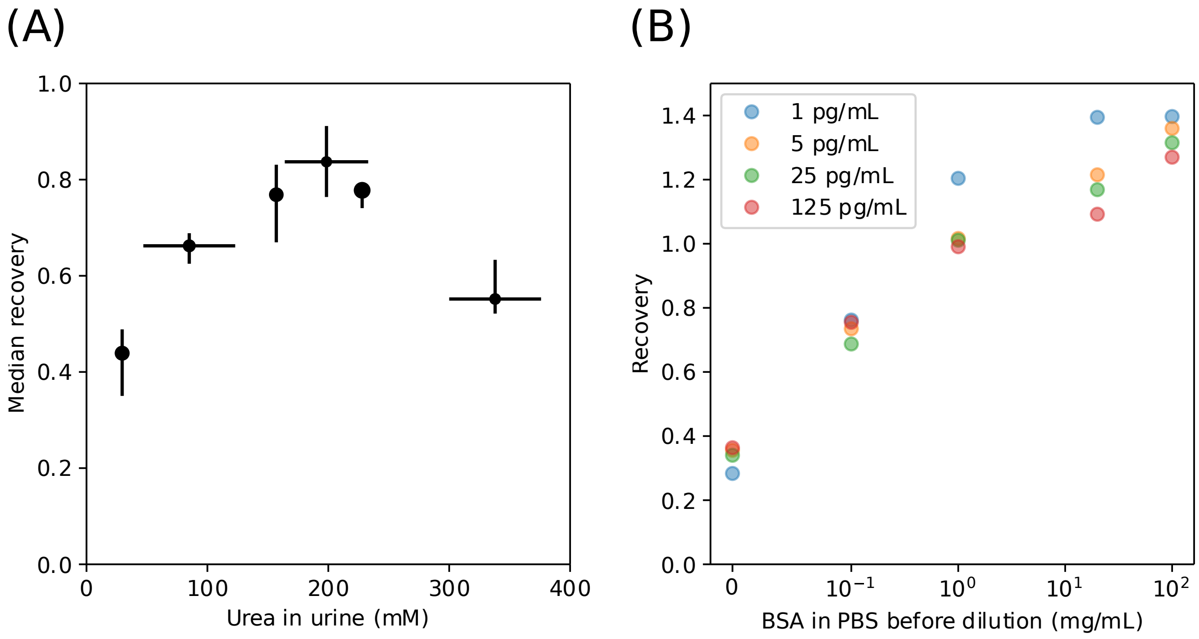


**Figure S6.** Recovery of Ag85B is negatively affected by urea in the urine and is dependent on urine total protein. (A) Urea concentrations in urine were measured using a Urea Nitrogen (BUN) Colorimetric Detection Kit (#EIABUN, Invitrogen). Recoveries were calculated as follows:

$$\% \, Recovery = 100\% \times \frac{Observed\ concentration - Endogenous\ concentration}{Spiked\ concentration}$$

. In the range of the urea tested, corresponding to physiological urea concentrations, reduced recovery was observed. (B) Different concentrations of Ag85B were spiked in PBS containing elevated concentrations of BSA. Without any BSA in the PBS to which Ag85B was spiked in, all spiked Ag85B concentrations resulted in poor recovery of ~ 35%.

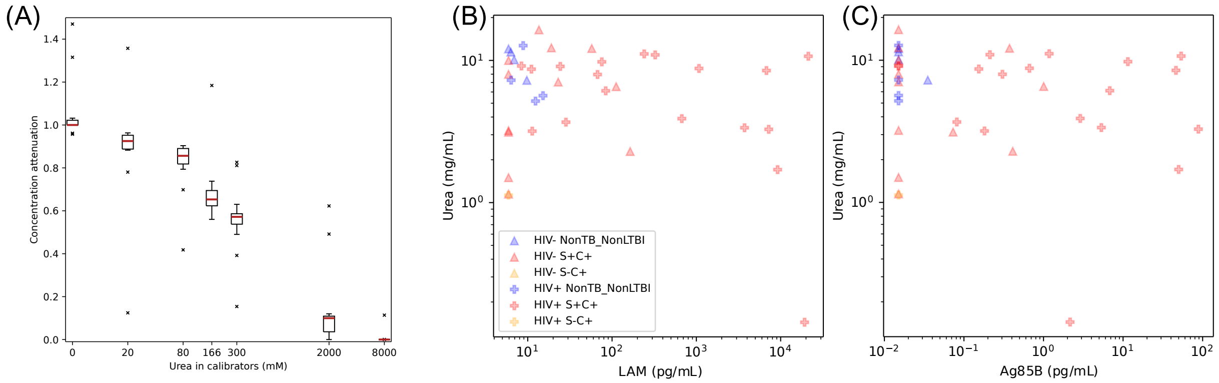


**Figure S7.** Urea attenuates Ag85B concentration and spans a wide range in urine samples. (A) Adding urea to the calibration curve attenuates Ag85B concentration. (B), (C) Urea concentrations in 40 samples from the training and validation cohorts, measured using a Urea Nitrogen (BUN) Colorimetric Detection Kit (#EIABUN, Invitrogen). Urea concentrations range between ~17–166 mM across all LAM and Ag85B concentrations.

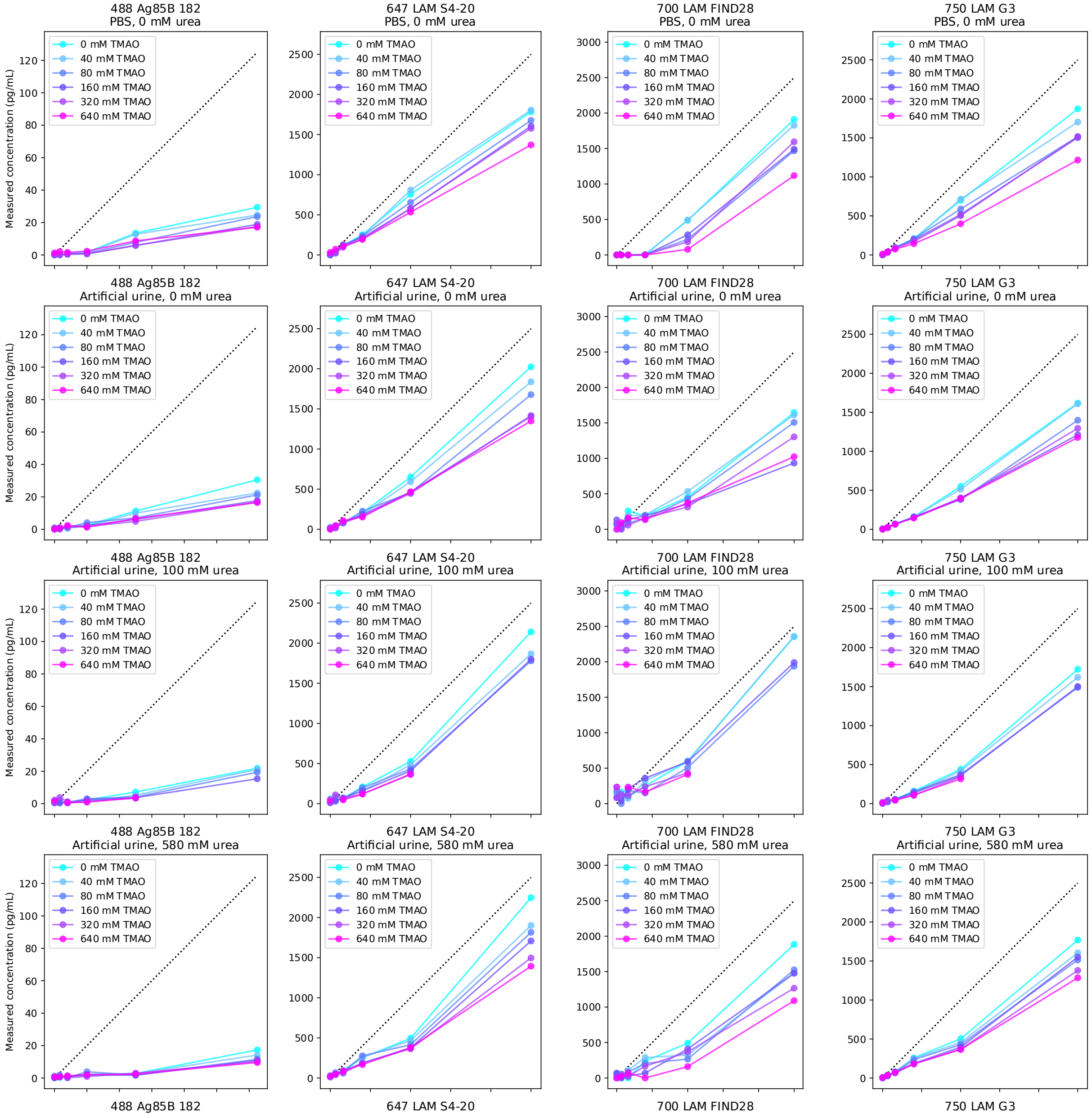


**Figure S8.** Attempts to improve recovery. Adding urea to the spiked matrix slightly improved the recovery of Ag85B. For example, 0 mM TMAO in 100 mM urea in artificial urine compared to 0 mM TMAO in 0 mM urea in artificial urine (light blue curve). The addition of TMAO did not yield higher recoveries. Lower recoveries are seen at the highest TMAO concentrations.

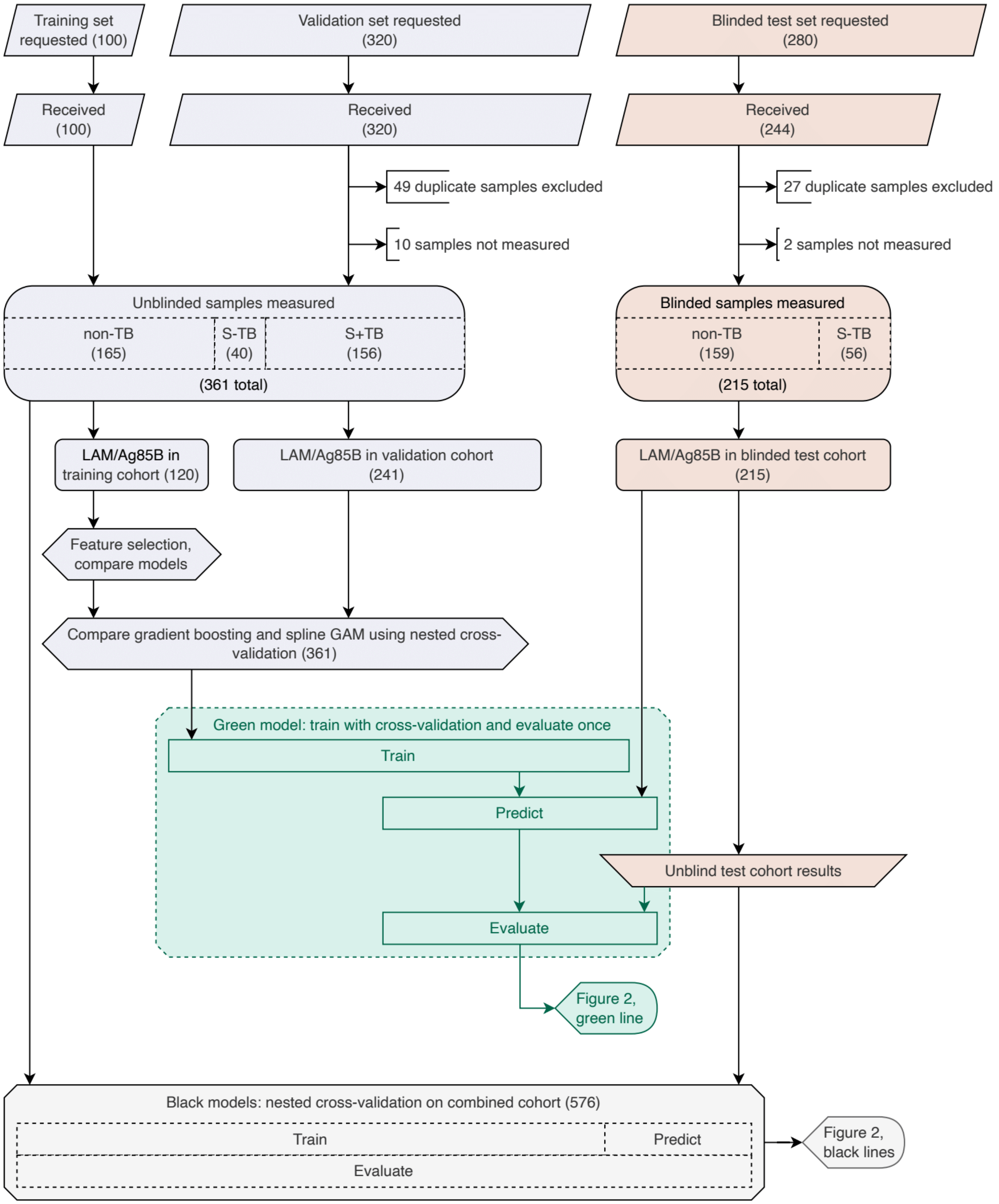


**Figure S9.** Flow diagram of model training and evaluation.

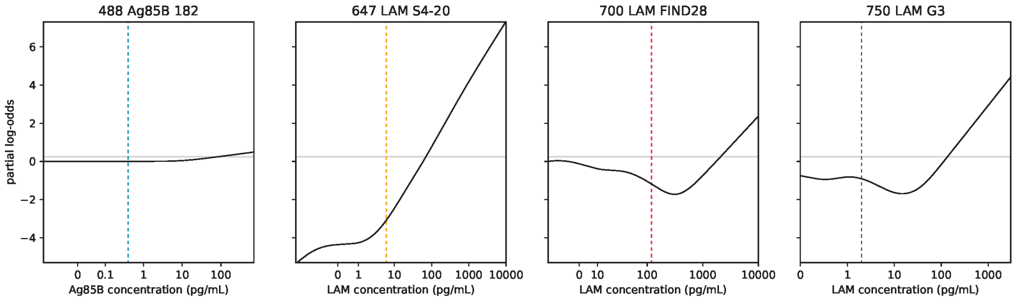


**Figure S10.** Partial dependence plots for the model evaluated on the test cohort. Both axes are arbitrary units due to transformations. Each of these four plots consists of a univariate spline model, and the log-odds assigned to a sample is the sum of all four. The predicted probability score between 0 and 1 is related to this log-odds by a logistic link function.

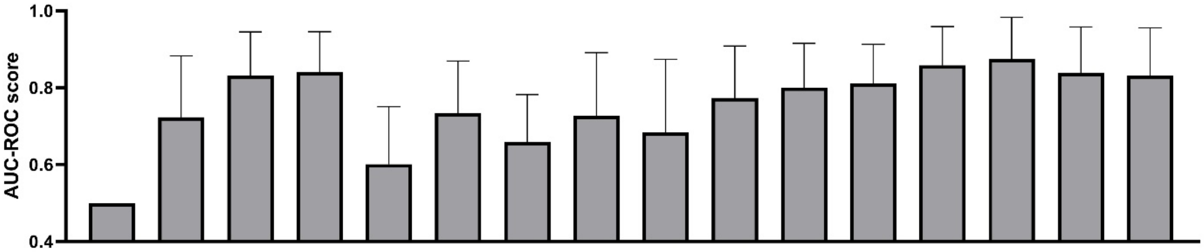


| | | | | | | | | | | | | | | | | |
|---|---|---|---|---|---|---|---|---|---|---|---|---|---|---|---|---|
| Ag85B | - | + | - | + | - | + | - | + | - | + | - | + | - | + | - | + |
| S4-20 | - | - | + | + | - | - | - | - | - | - | + | + | + | + | + | + |
| FIND28 | - | - | - | - | + | + | - | - | + | + | - | - | + | + | + | + |
| G3 | - | - | - | - | - | - | + | + | + | + | + | + | - | - | + | + |

**Figure S11.** AUC-ROC scores with different combinations of biomarkers. The AUC-ROC scores for each individual biomarker and different combinations were calculated. Adding Ag85B to all other combinations improved the AUC-ROC scores.

**Table S1**. Characteristics of common TB diagnostic tests

| | Sample type | Assay time | Time to result | Specificity | Sensitivity, S+HIV-PTB | Sensitivity, S-HIV-PTB | Sensitivity, HIV+PTB | Current usage |
|---|---|---|---|---|---|---|---|---|
| **Clinical** | Signs and symptoms | 10 minutes | <30 min | Variable | Variable | Variable | Variable | Widespread |
| **Radiography** | Chest X-ray | 15 minutes | <30 min | Variable | Variable | Variable | Variable | Used in some contexts |
| **Smear microscopy** | Sputum | 20 minutes | <30 min | 98% (N=5617) (49) | 100% (by definition) | 0% (by definition) | 34% (N=1339) (50) | Widespread |
| **AlereLAM** | Urine | 25 minutes | <30 min | 91% (N=2172) (2) | 16% (N=68) (51) | 2% (N=43) (51) | 42% (N=2172) (2) | Used in HIV+ contexts |
| **Truenat MTB** | Sputum | 1 hours | 1–3 h | 98% (N=1093) (14) | 91% (N=177) (14) | 36% (N=86) (14) | Not tested | Beginning rollout |
| **Truenat Plus** | Sputum | 1 hours | 1–3 h | 96% (N=1093) (14) | 96% (N=177) (14) | 47% (N=86) (14) | Not tested | Beginning rollout |
| **Simoa** | Urine | 1 hours 18 minutes | 1–3 h | 98% (N=576) | 48% (N=91) | 13% (N=39) | 45% (N=576) | Lab research only |
| **Xpert MTB/RIF** | Sputum | 2 hours | 1–3 h | 98% (N=598) (22) | 99% (N=598) (22) | 61% (N=379) (22) | 75% (N=635) (22) | Increasingly widespread |
| **Xpert Ultra** | Sputum | 2 hours | 1–3 h | 96% (N=1851) (22) | 99% (N=593) (22) | 78% (N=378) (22) | 88% (627) (22) | Increasingly widespread |
| **EclLAM** | Urine | 2 hours 10 minutes | 1–3 h | 98% (N=261) (51) | 84% (N=68) (51) | 19% (N=43) (51) | Not tested | Lab research only |
| **Culture (liquid)** | Sputum | 13 | 2–4 weeks | | 100% (by definition) | | | Confirmatory |
| **Culture (solid)** | Sputum | 26 | 2–4 weeks | | 100% (by definition) | | | Confirmatory |

**Table S2.** Alere LAM results using 244 samples.

| | Alere LAM | FIND |
|---|---|---|
| **Positive** | 22 | 58 |
| **Negative** | 222 | 186 |
| **TP** | 7 | |
| **FN** | 51 | |
| **TN** | 171 | |
| **FP** | 15 | |
| **Sensitivity** | 12% | |
| **Specificity** | 91.9% | |

**Table S3. TB biomarker Simoa assay characteristics.** LOD: limit of detection

| Marker | LOD (pg/mL) | Average Recovery % (min-max) | Average Dilution Linearity Fitted $R^2$ |
|---|---|---|---|
| Ag85A | 0.017 | 107 (92-130) | >0.99 |
| Ag85B | 0.015 | 109 (86-146) | 0.99 |
| Ag85C | 0.001 | 114 (95-138) | >0.99 |
| Adk | 0.16 | 111 (70-175) | 0.99 |
| GroES | 0.015 | 101 (75-141) | 0.98 |
| GlcB | 0.025 | 109 (82-135) | >0.99 |
| RelG | 0.004 | 119 (99-168) | >0.99 |
| LAM | 5.91 | 90 (70-113) | 0.98 |
| CFP10 | 0.56 | 86 (52-155) | 0.98 |
| Mce1A | 0.70 | 94 (36-162) | 0.94 |
| HspX | 0.069 | 88 (19-162) | >0.99 |

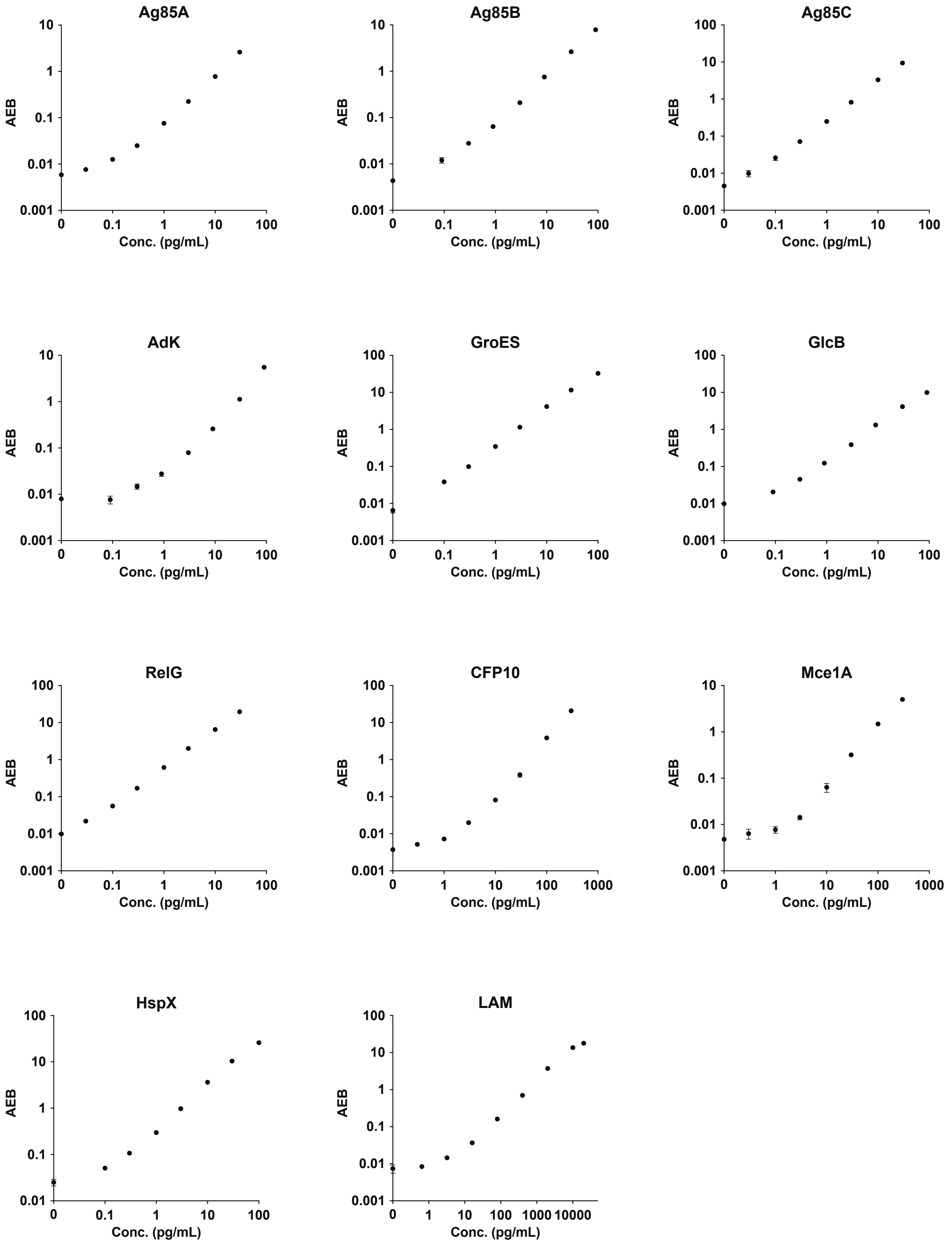


**Figure S12.** Calibration curves of all investigated markers. AEB is the signal unit for Simoa measurements. AEB: average enzyme per bead.

**Table S4.** Detectability of 11 TB markers in the discovery cohort. Data are n (%). Mann-Whitney test was used to test the significance of the difference between TB and non-TB patients with Bonferroni corrections. Undetectable samples were assigned a value at the assay limit of detection (LOD) to be included in the analysis.

| Antigen | LOD (pg/mL) | Number of detectable samples | | p value | Include/Exclude for model construction |
|---|---|---|---|---|---|
| | | TB (n=55) | Non-TB (n=45) | | |
| LAM | 5.91 | 53 (96%) | 13 (29%) | <0.0001 | Include |
| Ag85B | 0.015 | 45 (82%) | 19 (42%) | <0.0001 | Include |
| Ag85C | 0.001 | 37 (67%) | 22 (49%) | 0.094 | Exclude |
| Ag85A | 0.017 | 13 (24%) | 22 (49%) | - | Exclude |
| Adk | 0.16 | 9 (16%) | 13 (29%) | - | Exclude |
| GroES | 0.015 | 6 (11%) | 4 (9%) | - | Exclude |
| GlcB | 0.025 | 2 (4%) | 6 (13%) | - | Exclude |
| RelG | 0.004 | 5 (9%) | 8 (18%) | - | Exclude |
| CFP10 | 0.56 | 15 (27%) | 7 (16%) | - | Exclude |
| Mce1A | 0.70 | 6 (11%) | 9 (20%) | - | Exclude |
| HspX | 0.069 | 1 (2%) | 1 (2%) | - | Exclude |

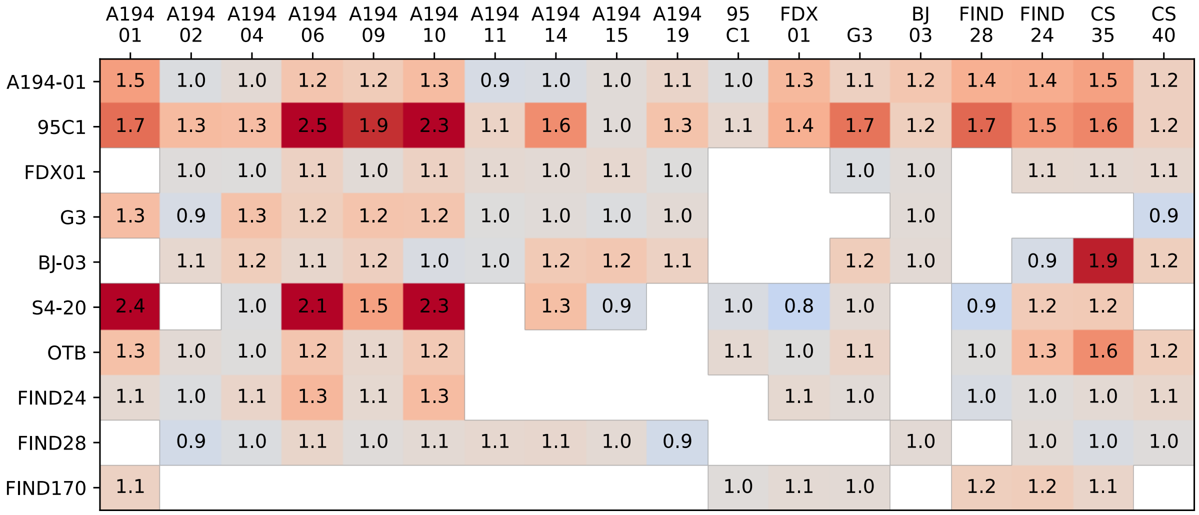


**Figure S13**. Cross-testing LAM antibodies.
